# The decline of COVID-19 severity and lethality over two years of pandemic

**DOI:** 10.1101/2022.07.01.22277137

**Authors:** Valentina Marziano, Giorgio Guzzetta, Francesco Menegale, Chiara Sacco, Daniele Petrone, Alberto Mateo Urdiales, Martina Del Manso, Antonino Bella, Massimo Fabiani, Maria Fenicia Vescio, Flavia Riccardo, Piero Poletti, Mattia Manica, Agnese Zardini, Valeria d’Andrea, Filippo Trentini, Paola Stefanelli, Giovanni Rezza, Anna Teresa Palamara, Silvio Brusaferro, Marco Ajelli, Patrizio Pezzotti, Stefano Merler

## Abstract

Undernotification of SARS-CoV-2 infections has been a major obstacle to the tracking of critical quantities such as infection attack rates and the probability of severe and lethal outcomes. We use a model of SARS-CoV-2 transmission and vaccination informed by epidemiological and genomic surveillance data to estimate the number of daily infections occurred in Italy in the first two years of pandemic. We estimate that the attack rate of ancestral lineages, Alpha, and Delta were in a similar range (10-17%, range of 95% CI: 7-23%), while that of Omicron until February 20, 2022, was remarkably higher (51%, 95%CI: 33-70%). The combined effect of vaccination, immunity from natural infection, change in variant features, and improved patient management massively reduced the probabilities of hospitalization, admission to intensive care, and death given infection, with 20 to 40-fold reductions during the period of dominance of Omicron compared to the initial acute phase.

## Introduction

The first two years of the COVID-19 pandemic have been characterized by an ever-changing epidemiological situation, forcing almost every country in the world to face a series of major challenges (1). During the first pandemic year, non-pharmaceutical interventions (NPIs) were widely adopted to counter the spread of COVID-19 and prevent health care systems to be overwhelmed, including social distancing restrictions culminating in nation-wide lockdowns, school closures and mandatory face masks (2–6). In Europe, the second pandemic year was characterized by a progressive relaxation of restrictions, the rollout of COVID-19 vaccination campaigns (7), and, concurrently, by the emergence of hyper transmissible SARS-CoV-2 variants of concern (8–11).

One of the main obstacles to pandemic containment has been represented by underreported and underdiagnosed SARS-CoV-2 infections. Due to the unknown extent of unobserved SARS-CoV-2 transmission, many aspects of the temporal changes in the COVID-19 epidemiology over the course of the pandemic remain unclear. Infection ascertainment ratios likely changed over time due to several factors such as the improvement in testing capacity, the increasing availability of diagnostics tests (also sustained by the development of quicker and cheaper antigen-based detection technologies), the varying intensity of contact-tracing, the shift of infections towards segments of the population less likely to develop symptoms, differences in pathogenicity associated with SARS-CoV-2 variants, the impact of external regulations (e.g. the requirement of a negative test result for access to workplaces or community indoor spaces in absence of vaccination), and changes in the people’s attitudes and behavior related to SARS-CoV-2 testing or in the self-perception of symptoms associated with COVID-19. The quantification of changes in population susceptibility to SARS-CoV-2 infection is instrumental to assess the expected impact of further epidemic waves. This has been a difficult task due to the lack of knowledge on the number of undetected infections throughout the pandemic; to the partial protection against infection provided by existing vaccines (12–14); to the emergence of new variants partially escaping immunity from natural infection with previous lineages (15); and to the effect of waning of protection from both natural infection and vaccination (12–14,16). If the actual number of SARS-CoV-2 infections is unknown, it is also not possible to estimate changes in the proportion of infections resulting in adverse outcomes (e.g., severe disease or death). In turn, these changes were induced by COVID-19 vaccines, which have proven highly effective in preventing severe COVID-19 or death (12,17,18), as well as by the improvement in timely diagnoses and clinical care, by the increased health care capacity, and possibly also by changes in virulence of emerging SARS-COV-2 variants (19,20). An assessment of the immunity accrued in the first two pandemic years and of the probability of adverse outcomes is essential to quantify the risks associated to SARS-CoV-2 infection and COVID-19 burden in the future.

In this study, we use a mathematical model of SARS-CoV-2 transmission, informed with integrated surveillance data (epidemiological and microbiological results on variants), to quantify the evolution of COVID-19 epidemiology in Italy over the first two years of the pandemic. Specifically, we provide estimates of changes in infection ascertainment ratios, infection attack rates, population susceptibility to infection, and probability of adverse outcomes given SARS-CoV-2 infection.

## Results

We developed an age-structured stochastic model to simulate SARS-CoV-2 transmission and vaccination in Italy between February 21, 2020, (when the first locally transmitted case was detected) and February 20, 2022. We divided the two years of simulation into five phases (background colors in Figure 1a). The first two phases are associated with the circulation of ancestral SARS-CoV-2 lineages and they distinguish the first pandemic wave including the national lockdown (phase 1, from February 21, 2020 to the end of June 2020), and a second phase characterized by a new upsurge of cases in fall 2020 and by the start of the COVID-19 vaccination campaign on December 27, 2020 (phase 2, from July 1, 2020 to February 17, 2021). The three remaining phases correspond to the periods of dominance in Italy of different SARS-CoV-2 variants: Alpha (phase 3, from February 18, 2021 to July 1, 2021), Delta (phase 4, from July 2, 2021 to December 23, 2021), and Omicron (phase 5, from December 24, 2021 to February 20, 2022) (21). The epidemic curve over the study period (Figure 1a) was reproduced by adjusting the SARS-CoV-2 transmission rate on a given day in such a way that the model reproduction number matches the daily net reproduction number Rt as estimated from the epidemiological surveillance data (29,30). The model keeps into account the dynamics of age-specific population immunity due to natural infection, to the progress of the vaccination campaign including first, second and booster doses (Figure 1b) (22). The model considers waning of immunity after natural infection with all lineages (16), as well as the ability of the omicron variant to escape immune response from natural infection with previous SARS-CoV-2 lineages (15).Vaccine protection is assumed to be “leaky”, i.e. successfully vaccinated individuals are partially immune with a relative risk of infection that depends on the SARS-CoV-2 variant and on the number of doses received and on the time since vaccination to reproduce the effect of waning immunity (12–14). Further details on the model are provided in Section Methods and in Appendix.

**Figure 1.**
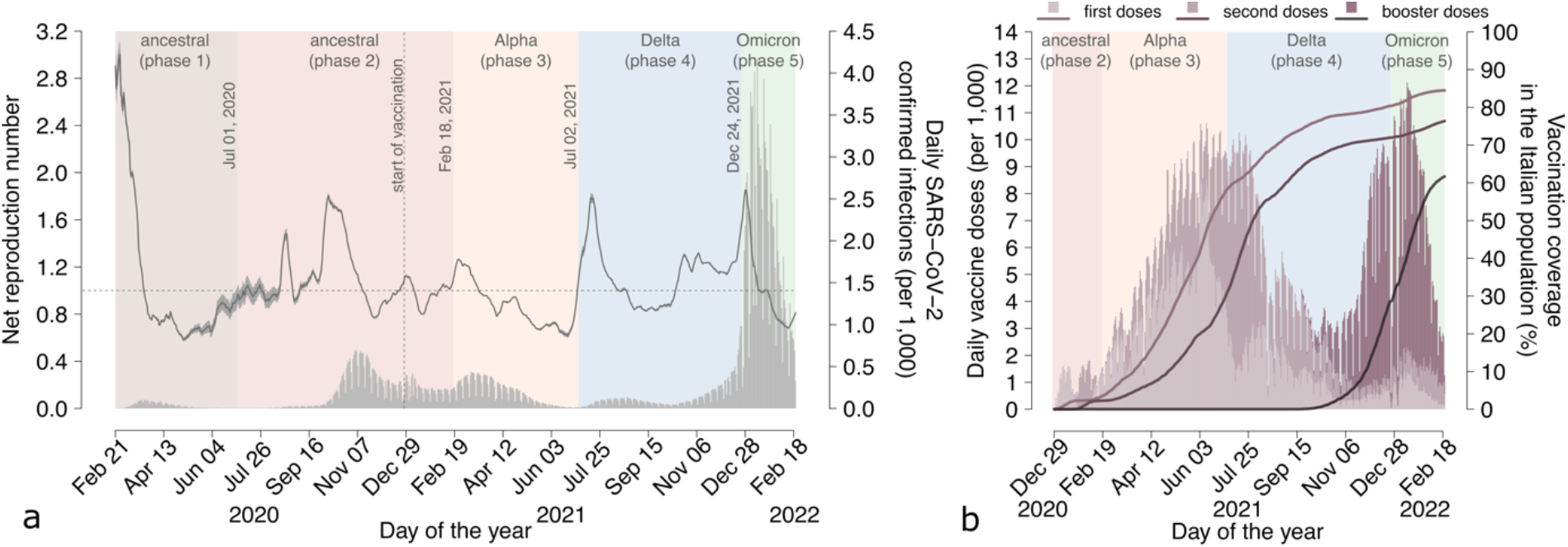
Epidemiological profile of the SARS-CoV-2 epidemic in Italy. **a** Mean estimates of the net reproduction number Rt as obtained from epidemic curves of symptomatic cases by date of symptom onset collected by the National Integrated Surveillance System (23) (mean, grey solid line; shaded area, 95% CI; y-axis on the left). Horizontal dotted line: epidemic threshold (Rt = 1). Grey bars represent the daily incidence per 1,000 individuals of SARS-CoV-2 confirmed infections by date of diagnosis as reported to the Italian Integrated Surveillance System (24,25) (y-axis on the right). Background colors indicate the classification in different phases, and the dates indicated within the graph denote the day of transition between consecutive phases. The vertical dotted line denotes the start of the vaccination campaign on December 27, 2020. **b** Daily number of vaccine doses administered in Italy per 1,000 individuals (stacked barchart, y-axis on the left) (22). Line and bar colors, from lighter to darker shades, indicate respectively first, second and booster doses. Solid lines show the cumulative vaccination coverage in the Italian population (y-axis on the right). In Italy, administration of two doses is recommended to all individuals aged 5 years or more; administration of one booster dose is recommended to all individuals aged 12 years or more.

### Attack rate and ascertainment of SARS-CoV-2 infections

Despite the explosive spread of SARS-CoV-2 in the early phase of the pandemic, which threatened to overwhelm the Italian health system, we estimate that the adoption of a strict nationwide lockdown managed to limit the infection attack rate in the first phase to 2.8% (95%CI: 1.8-3.6) (Figure 2a). During the second phase, in the context of less stringent NPIs, we estimate an attack rate of 11.4% (95%CI: 7.3-15.2). In both phases dominated by ancestral lineages, the SARS-CoV-2 attack rate was substantially homogeneous across age groups (Figure 2a). We estimate that in the first phase, about 15.0% (95%CI: 11.1-22.5) of infections, i.e. about 1 in 7, were detected by the Italian Integrated Surveillance System (Figure 2b). Even though the second phase was characterized by a greater infection incidence, we estimate a higher infection ascertainment ratio of 38.6% (95%CI: 28.1-58.5) (Figure 2b).

**Figure 2.**
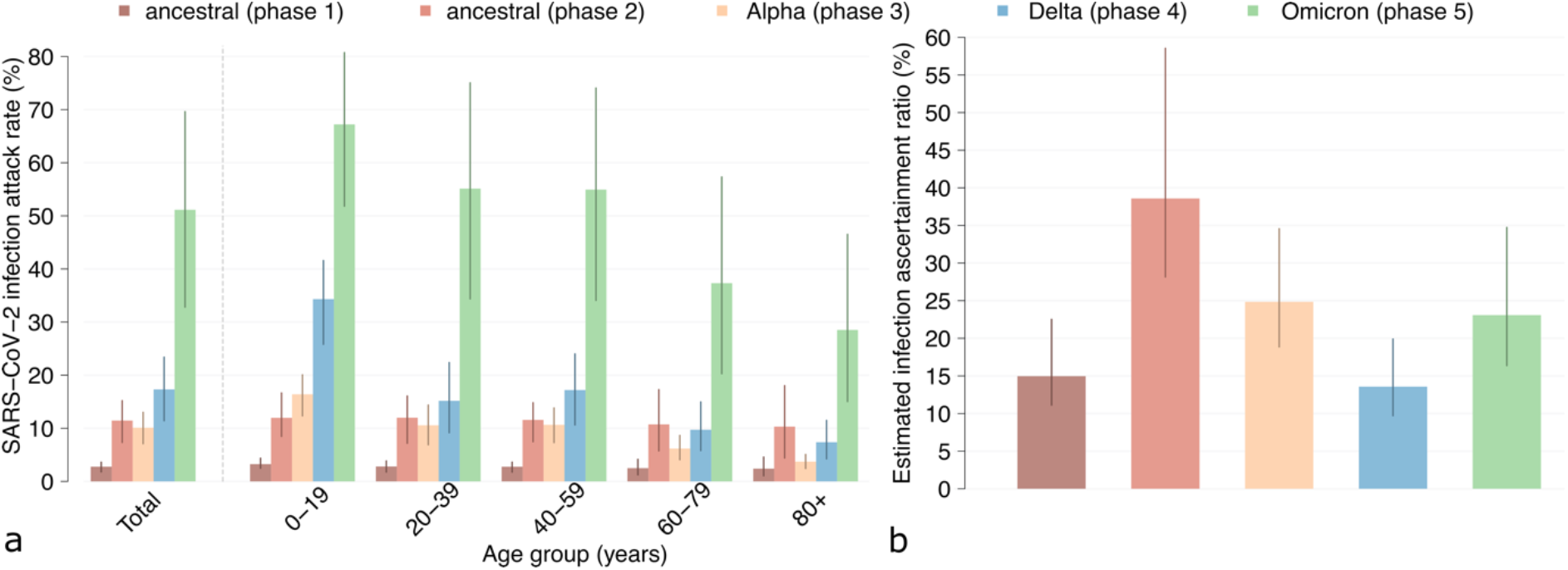
SARS-CoV-2 attack rates and infection ascertainment ratios. **a** Estimated phase-specific SARS-CoV-2 infection attack rate (%) as of February 20, 2022, in the overall population and by age classes. Colors indicate the considered phase. Bars: mean estimates; vertical lines: 95% CI; n= 300 stochastic model realizations. **b** Estimated SARS-CoV-2 infection ascertainment ratio in the different phases (%). Bars: mean estimates; vertical lines: 95% CI; n= 300 stochastic model realizations. The infection ascertainment ratio in each phase is estimated as the fraction between the number of SARS-CoV-2 confirmed infections as reported to the Italian Integrated Surveillance System and the number of infections estimated through the model in each phase.

By mid-February 2021, the ancestral SARS-CoV-2 lineages were replaced by the more transmissible Alpha variant (8) which remained dominant until early July 2021. During this period, we estimate that Alpha infected about 10.1% (95%CI: 7.1-13.0) of the Italian population, with a marked heterogeneity across ages: the highest attack rate is estimated in the age group 0-19 years (16.4%, 95%CI: 12.3-20.1), while the lowest in people aged over 80 years (3.7%, 95%CI: 2.4-5.1) who had been prioritized for vaccination in the early months of 2021 (Figure 2a and Figure S2). We estimate for this period an infection ascertainment ratio of 24.9% (95%CI: 18.9-34.6) (Figure 2b).

The second half of 2021 was characterized by the circulation of the Delta variant, in the context of a progressive relaxation of NPIs, with an estimated attack rate of 17.3% (95%CI: 11.4-23.4). Our results suggest that the progression of the vaccination campaign, including the administration of booster doses (Figure 1b), led to a further shift of infections towards less protected age groups (children and young adults), with over one third of infections occurring among individuals aged 20 years or less (Figure S4). For this period, we estimate a further decrease in the infection ascertainment ratio with respect to the Alpha phase, 13.6% (95%CI: 9.7-19.9) (Figure 2b).

By the end of December 2021, the Delta variant was replaced by Omicron. We estimate that, as of February 20, 2022, about 51.1% (95%CI: 32.8-69.6) of the Italian population got infected with Omicron, with age-specific attack rates ranging from 28.5% (95%CI: 15.0-46.5) in the individuals aged 80 years or more to 67.2% (95%CI: 51.8-80.7) in those younger than 20 years. For this period, we estimate an infection ascertainment ratio of 23.1% (95%CI: 16.4-34.7).

### Evolution of population susceptibility

Throughout the first two years of the COVID-19 pandemic, the percentage of the population susceptible to SARS-CoV-2 infection (depicted in shades of grey in Figure 3a) progressively decreased from 97.5% (95%CI 96.8-98.4) at the end of the first phase to 13.0% (95%CI: 5.1-23.5) by February 20, 2022. The effect of vaccination on population susceptibility was still negligible when the ancestral strain was replaced by the Alpha variant, with only 2.9% (95%CI: 2.8-3.0) of the population being protected by vaccination (orange in Figure 3a and 3c), while it had become significant at the end of the Alpha phase (Figure 3d): according to our estimates, only about one third of the population (33.0%, 95%CI: 30.4-35.6) was unprotected against SARS-CoV-2 infection by July 2021, most of them being children and young adults (Figure 3a and 3d).

**Figure 3.**
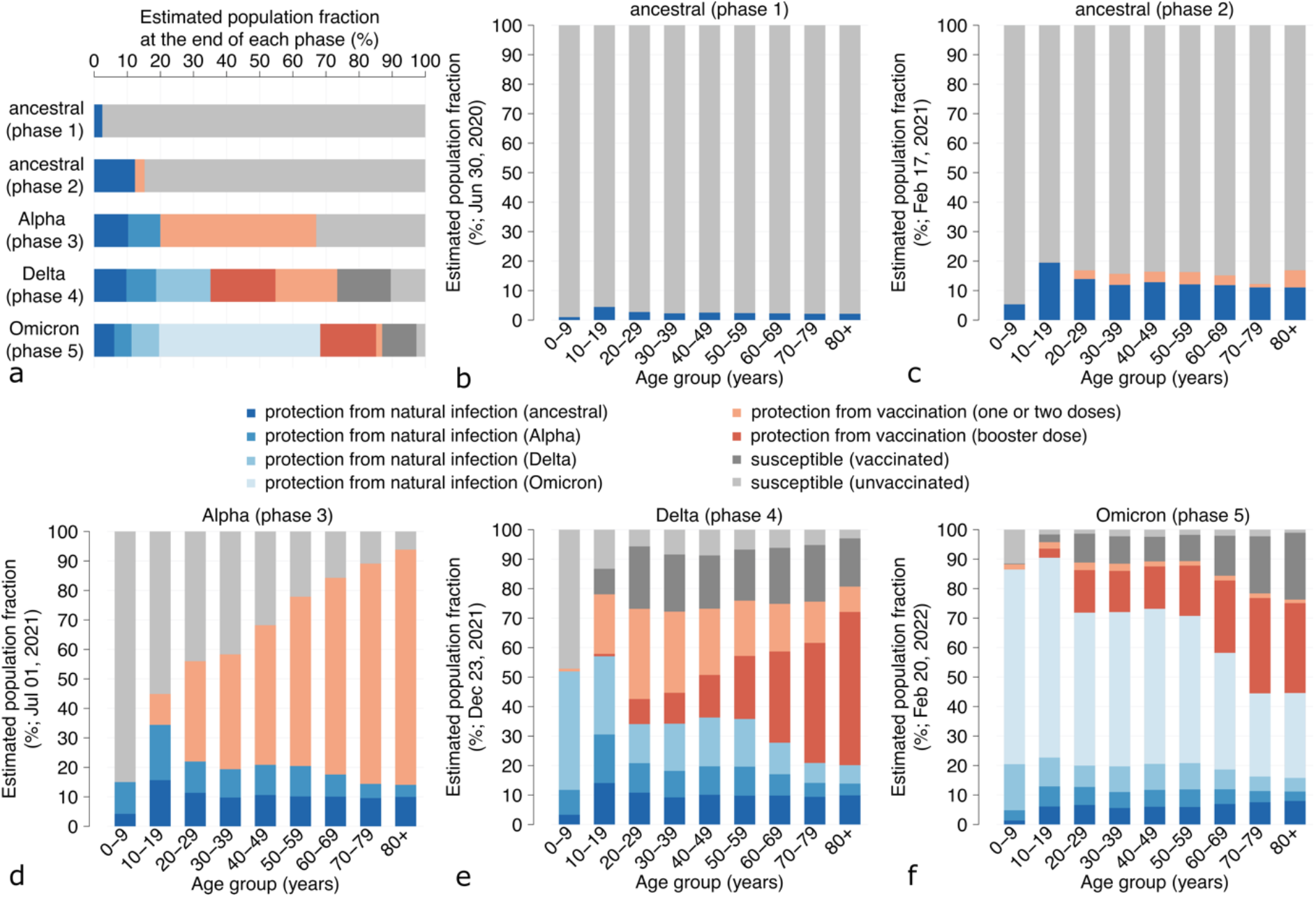
SARS-CoV-2 susceptibility profiles. **a** Mean estimates for the overall SARS-CoV-2 susceptibility profile of the Italian population at the end of each phase (n= 300 stochastic model realizations). In categories “protection from natural infection” the indicated variant corresponds to that of the most recent infection, independently of whether the individual has been vaccinated before or after natural infection. Uninfected and vaccinated individuals whose immunity has not waned are included in the “protection from vaccination” categories, depending on whether they received or not a booster dose. Individuals who were never infected or whose immunity against infection (from natural infection or vaccine) has waned are considered in the “susceptible” categories depending on their vaccination status. Note that the latter may retain a residual protection against other clinical endpoints, such as symptoms, severe disease, or death. Also note that individuals who experienced SARS-CoV-2 infection (depicted in shades of blue) are considered fully protected against reinfection during the ancestral, Alpha and Delta phases, while in the Omicron phase they may get infected due to the significant ability of this variant to escape previous immunity from natural infection. **b** Mean estimates of the SARS-CoV-2 susceptibility profile by age groups at the end of the first ancestral phase (June 30, 2020). **c** As **b** but at the end of the second ancestral phase (February 17, 2021). **d** As **b** but at the end of the Alpha phase (July 1, 2021). **e** As **b** but at the end of the Delta phase (December 23, 2021). **f** As **b** but at the end of simulations (February 20, 2022, Omicron phase).

The Delta phase was characterized by an increased vaccination coverage in children and young adults and by the start of the booster dose campaign, which initially targeted the over 80 population and then was gradually extended to younger age groups (Figure 3e). The administration of boosters partially compensated the waning of immunity after two doses of vaccine. However, by the end of December 2021, when Delta was replaced by Omicron, individuals unprotected against SARS-CoV-2 infection still represented 26.6% (95%CI: 21.1-32.4) of the Italian population (Figure 3a).

We estimate that by February 20, 2022, 68.3% (95%CI: 48.6-84.3) of the Italian population was protected by natural immunity (blue tones in Figure 3a and 3f) mostly acquired after infection with Omicron (48.7%, 95%CI: 31.4-65.9). This percentage includes also individuals vaccinated before or after natural infection with SARS-CoV-2. Vaccination alone protected an additional 18.7% (95%CI: 10.7-27.9), mainly thanks to the administration of boosters (orange and red in Figure 3f). However, we estimate that by February 20, 2022, a marked proportion of individuals unprotected against SARS-CoV-2 infection can be found among vaccinated subjects (especially among the adults and elderly) due to the waning of vaccine protection (dark grey in Figure 3f). At that time, the contribution of unvaccinated individuals to residual population susceptibility was negligible (2.7% of the population, 95%CI: 0.9-5.7), with a large majority represented by children younger than 10 years.

### Evolution of COVID-19 severity and lethality

We estimated the probability of hospitalization, admission to ICU, and death after SARS-CoV-2 infection in the different epidemic phases (Figure 4, first row). The first ancestral phase was characterized by the highest probability of severe clinical outcomes, with a probability of hospitalization per infection of 5.4% (95%CI: 4.0-8.2), a probability of ICU admission of 0.65% (95%CI: 0.48-0.97) and a probability of death of 2.2% (95%CI: 1.7-3.4). Estimates of all these probabilities progressively decreased throughout the pandemic.

**Figure 4.**
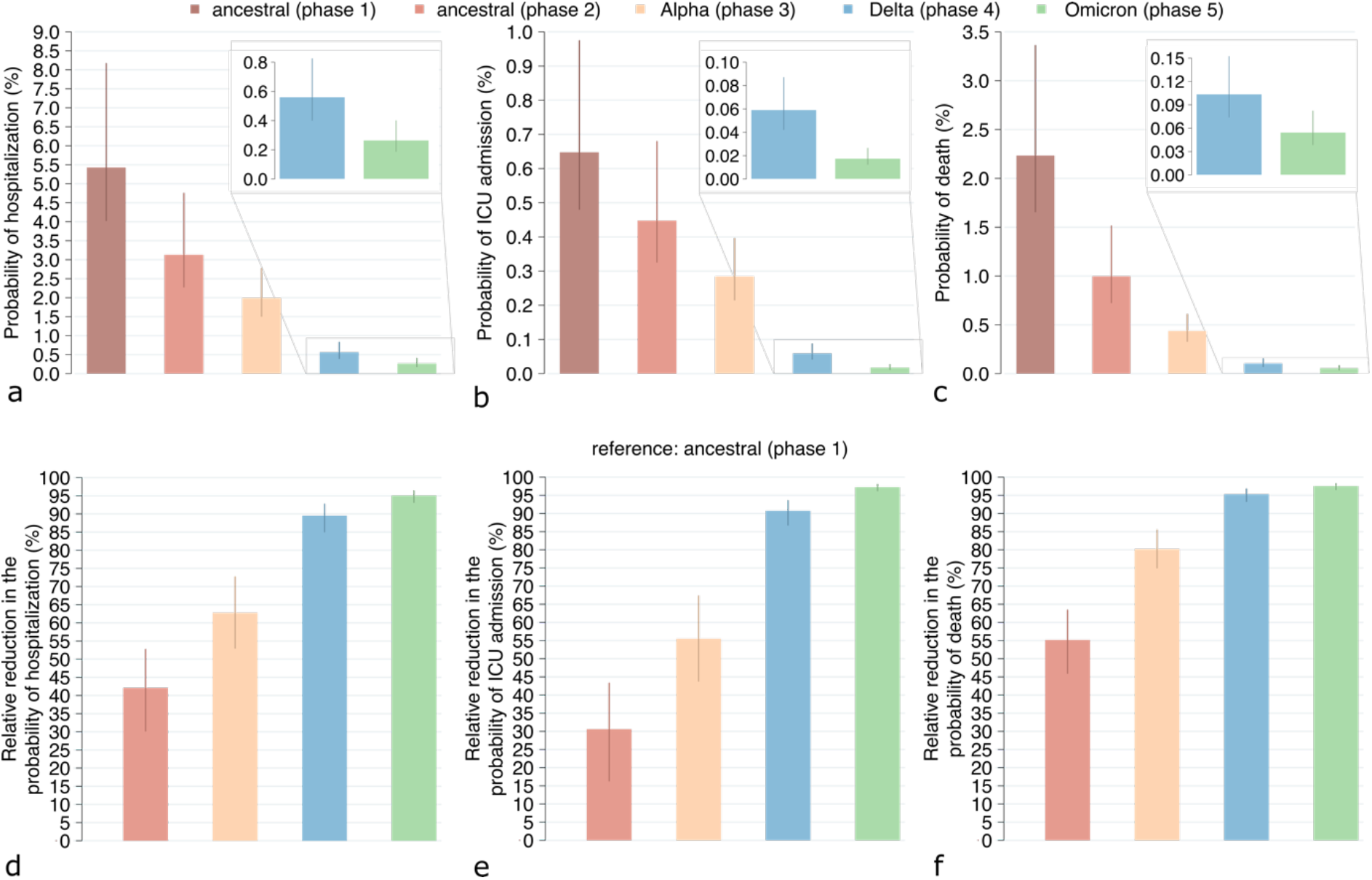
Probabilities of adverse outcomes given SARS-CoV-2 infection. a Probability of being hospitalized in the different epidemic phases (%), computed as the ratio between the number of COVID-19 hospital admissions reported to the Italian Integrated Surveillance System and the estimated number of SARS-CoV-2 infections in the same phase. b Probability of being admitted to an ICU in the different phases (%), computed as the ratio between the number of SARS-CoV-2 admissions to ICUs reported to the Italian Integrated Surveillance System and the estimated number of SARS-CoV-2 infections in the same phase. c Probability of death in the different phases (%), computed as the ratio between the number of COVID-19 deaths reported to the Italian Integrated Surveillance System and the estimated number of SARS-CoV-2 infections in the same phase. d Estimated relative reduction in the probability of hospitalization in the different phases compared to the first ancestral phase (%). e As d but for the probability of ICU admission. f As d but for the probability of death. Bars: mean estimates; vertical lines: 95% CI; n= 300 stochastic model realizations.

The probability of hospitalization given infection is estimated to decrease to 3.1% (95%CI: 2.3-4.7) in the second ancestral phase (42.1% reduction compared to first ancestral phase), 2.0% (95%CI: 1.5-2.8) in the Alpha phase (62.8% reduction compared to first ancestral phase; 35.8% compared to second ancestral phase), 0.56% (95%CI: 0.40-0.82) in the Delta phase (89.5% reduction compared to first ancestral phase; 71.9% compared to Alpha phase) and 0.27% (95%CI: 0.19-0.40) in the Omicron phase (95.1% reduction compared to first ancestral phase and 52.7% compared to Delta phase) (Figure 4a and 4d).

The probability of ICU admission given infection is estimated to decrease to 0.45% (95%CI: 0.33-0.68) in the second ancestral phase (30.6% reduction compared to first ancestral phase), to 0.28% (95%CI: 0.22-0.40) in the Alpha phase (55.5% reduction compared to first ancestral phase; 35.9% compared to second ancestral phase), to 0.06% (95%CI: 0.04-0.09) in the Delta phase (90.7% reduction compared to first ancestral phase and 79.2% compared to Alpha phase) and to 0.018% (95%CI: 0.012-0.026) in the Omicron phase (97.3% reduction compared to first ancestral phase and 70.3% compared to Delta phase) (Figure 4b and 4e).

Finally, we estimate the probability of death given infection at 1.0% (95%CI: 0.7-1.5) in the second ancestral phase (55.2% reduction compared to first ancestral phase), 0.44% (95%CI: 0.33-0.61) in the Alpha phase (80.2% reduction compared to the first ancestral phase; 55.9% compared to second ancestral phase), 0.1% (95%CI: 0.07-0.15) in the Delta phase (95.3% reduction compared to first ancestral phase; 76.3% compared to Alpha phase) and 0.05% (95%CI: 0.04-0.08) in the Omicron phase (97.5% reduction compared to first ancestral phase; 47.3% compared to Delta phase) (Figure 4c and 4f).

## Discussion

In this work, we analyzed the first two years of the COVID-19 pandemic in Italy and quantified changes in COVID-19 epidemiological indicators, including the infection ascertainment ratio, the infection attack rate, the population susceptibility to SARS-CoV-2 infection, and the probabilities of hospitalization, ICU admission, and death, given infection.

Detection of SARS-CoV-2 infections changed throughout the pandemic. We estimated an increase in the infection ascertainment ratio between the two ancestral phases, likely ascribable to the expanding of testing capacity (Figure S5), to the strengthening of regional reporting systems and to an aggressive implementation of the test-track and trace strategy. In contrast, we found a reduction in infection ascertainment during the Alpha and Delta phases, probably related to a combination of factors including: the availability of home testing leading to self-diagnoses that were not notified to the surveillance system, the reduction in the frequency of contact tracing since 2021, and the significant increase of asymptomatic infections following the expansion of the vaccination program. Indeed, vaccination brought a shift of infections towards younger age groups (Figure S4) and increased the proportion of breakthrough infections (Figure S6); both trends reduced the overall probability of having symptoms given an infection (26–28) and therefore the overall probability of test-seeking by unaware infected individuals. In the Omicron phase, we estimated that the infection ascertainment ratio increased again. This period was characterized by a substantial scale-up of COVID-19 testing capacity (Figure S5) associated to the escalation of cases, as well as by a surge in voluntary testing in preparation for gatherings during Christmas and New Year’s holidays.

We estimated that the SARS-CoV-2 infection attack rate was highest for the Delta (17%) and Omicron (51%) phases, despite the highest vaccination coverage in the population. The large number of infections in these periods may be ascribable to several factors, such as a possible decline in adherence to residual COVID-19 restrictions due to pandemic fatigue (29); the high transmissibility of these variants (8–10,30); the reduced efficacy of the vaccine in preventing infection by different viral variants (12–14); the increased risk of reinfection during the Omicron phase compared with previous epidemic phases (31); the progressive release of restrictions, sustained by a lower morbidity among vaccinated individuals (27,28) and by a reduced intrinsic severity of Omicron (19,20).

Our results underline the key role played by booster vaccination during the Omicron phase. We estimated that, by the end of February 2022, about 9 out of 10 individuals in Italy who were still protected by vaccination had received a booster dose (Figure 3a). Given the available evidence on the declining effectiveness of boosters over time (13,14), waning immunity will likely contribute to an increase in population susceptibility throughout 2022 (Figure S7).

Estimates obtained for the probabilities of hospitalization, ICU admission and death given infection in the first phase of the pandemic (5.4%, 0.65% and 2.2%, respectively) are in line with values reported in the literature (32–34). We found that the severity of SARS-CoV-2 infections has progressively declined throughout the pandemic, with the infection fatality ratio in 2022 falling close to the levels of 2009 H1N1 pandemic influenza (estimated at about 0.02% (35)). Compared to the first pandemic wave, we estimated a 20 times lower probability of hospitalization, a 36 times lower probability of ICU admission, and a 40 times lower probability of death during the Omicron phase. The estimated reduction in COVID-19 severity is attributable to a combination of factors. Improved knowledge on the pathogen and patient management, and the relieving of the pressure on the health care system allowed by the national lockdown likely reduced severity between the first and second ancestral phases. From the Alpha phase on, the vaccination program increased the population immunity against severe disease. The predominance of Omicron variant likely contributed to a reduced severity of the disease due to a decrease in the intrinsic viral pathogenicity (19,20). In the absence of immunity from natural infection and vaccination the health impact of SARS-CoV-2 variants could have been different. We note however that, as of June 2022, the overall COVID-19 morbidity and mortality is still remarkably high in Italy due to the very high incidence of infections (25). For what concerns the estimated probability of ICU admission given infection, we note that this does not necessarily represent the probability of critical disease but includes the effect of patient management choices concerning trade-offs between the usage of limited ICU resources and the expected benefits for the patient; the expansion of ICU capacity throughout the pandemic may have influenced temporal changes in this parameter.

The proposed model was designed to evaluate overall changes of several COVID-19 epidemiological indicators at the national level, but it cannot estimate the relative weight of individual determinants in the reduction of the severity of SARS-CoV-2 infections. Estimates for COVID-19 severity rely on the assumption of negligible underdiagnosis of COVID-19 hospitalizations, admission to ICUs, and deaths. Another limitation of our analysis is that we consider conventionally defined epidemic phases with instantaneous transitions, roughly corresponding to the times at which different variants became dominant (Table S1). The susceptibility profile of the population is also updated instantaneously at every change of variant, to consider different vaccination effectiveness and rates of waning. This approximation is in contrast with the observation that SARS-CoV-2 variants showed extended periods of co-circulation (21), which are not explicitly modeled. Furthermore, we could not consider changes in the age-specific proportion of contacts over time in absence of longitudinal contact patterns data by age. This is obviously a simplification as, for example, some restrictions targeted preferentially contacts in specific age groups (e.g., school closure). Despite these conservative assumptions, we show that the model approximates well age-specific trends in SARS-CoV-2 infection dynamics (Figure S4).

Quantitative estimates provided in this study apply to the case of Italy and depend on many country-specific factors, such as governmental choices on mitigation measures, the speed of rollout of COVID-19 vaccines, or the population socio-demographic structure. Therefore, generalization to different geographic contexts and conditions should be made with caution. Nonetheless, we expect that the general trends and conclusions may apply to other high-income countries that have adopted a similar mitigation approach throughout the pandemic. In particular, considering that the Italian demographic structure is skewed towards older ages, the decreasing trends in probabilities of adverse outcomes might be even more marked in countries with younger populations.

Despite the large number of SARS-CoV-2 cases since the beginning of 2022, the burden of COVID-19 in Italy has remained limited with a manageable impact on hospitals. However, the possible future emergence of new variants that may escape previous immunity (natural or from vaccine), be more transmissible and/or pathogenic stresses the need of maintaining careful surveillance on SARS-CoV-2 variants (36) and epidemic trends. Moreover, our results highlight the key role played by the changing immunity against SARS-CoV-2 from natural infections and vaccination on the decrease of severe clinical outcomes upon SARS-CoV-2 infection across the different phases. This trend might be reversed if the level of population susceptibility to SARS-CoV-2 infection will increase again in the future, due to waning of vaccine and natural protection.

## Methods

We developed an age-structured stochastic model of SARS-CoV-2 transmission and vaccination, based on a susceptible-infectious-removed-susceptible (SIRS) scheme (37,38). The model was used to simulate the evolution of COVID-19 epidemiology in Italy between February 21, 2020, and February 20, 2022. The simulation period is conventionally subdivided as follows:

1. “ancestral (phase 1)”, from February 21 to June 30, 2020.
2. “ancestral (phase 2)”, from July 1, 2020, to February 17, 2021.
3. “Alpha (phase 3)”, from February 18 to July 1, 2021.
4. “Delta (phase 4)”, from July 2 to December 23, 2021.
5. “Omicron (phase 5)”, from December 24, 2021 to February 20, 2022.

Dates of transition between variants were chosen based on estimates of the prevalence of SARS-CoV-2 lineages from genomic surveillance data in Italy (Table S1) (21). The model population is stratified by age (17 5-year age groups from 0 to 84 years plus one age group for individuals aged 85 years or older). Mixing patterns across ages are encoded by a social contact matrix estimated prior to the COVID-19 pandemic (39). During circulation of ancestral lineages susceptibility to SARS-CoV-2 infection was assumed to be age-dependent (lower in children under 15 years of age and higher for the elderly above 65 years, compared to individuals aged 15 to 64 years) (40). For SARS-CoV-2 variants, we assumed homogenous susceptibility across ages in absence of age-specific estimates. Infectiousness was assumed to be homogeneous by age for all lineages (40,41).

To reproduce the epidemic curve over the study period, we adjusted the SARS-CoV-2 transmission rate on a given day, in such a way that the model’s reproduction number (estimated via the Next Generation Matrix approach (42,43)) would match the daily net reproduction number Rt as estimated from surveillance data (specifically, the number of new symptomatic SARS-CoV-2 infected individuals by date of symptom onset) (29,30).

The rollout of the vaccination campaign is modeled using detailed data on the daily age-specific number of doses administered over the considered period, including first, second and booster doses. In the model, individuals are considered eligible for vaccination, independently of a previous SARS-CoV-2 infection. Breakthrough infections (i.e., infections in vaccinated individuals) are assumed to be half as infectious as those in unvaccinated individuals (44,45).

Protection from natural immune response is assumed to wane exponentially with a constant rate over all periods considered (16). Before waning, natural infection provides complete protection against re-infection with ancestral lineages and Alpha and Delta variants, while we assume a partial cross-protection against Omicron (15). Waning of protection from vaccine-induced immune response was observed only in the Delta and Omicron phases (12–14). Accordingly, we consider variant-specific average durations of protection after two doses of vaccine and after a booster dose. Before waning of vaccine-induced immunity, individuals are in a compartment where the risk of infection is reduced by the vaccine efficacy. After waning of immunity either from natural infection or vaccination, individuals are in a compartment where the risk of infection is the same as unvaccinated individuals who were never exposed to SARS-CoV-2. We note that individuals with waned protection against infection may still be protected against other clinical endpoints, such as symptoms, severe disease, or death (12).

The age-profile of SARS-CoV-2 susceptibility in the Italian population at the end of each phase is obtained from model states variables, considering the protection acquired from natural infection or vaccination and accounting for waning of both. The SARS-CoV-2 infection ascertainment ratio is computed as the ratio between COVID-19 cases reported to the Italian Integrated Surveillance System (24) and the number of SARS-CoV-2 infections estimated by the model in the same period. The probabilities of hospitalization, ICU admission and death after SARS-CoV-2 infection are computed as the ratio between COVID-19 cases reported to the Italian Integrated Surveillance System (24) who were respectively admitted to hospital, ICUs, and deceased, and the number of SARS-CoV-2 infections estimated by the model in the same period. COVID-19 cases are assigned to each period based on their date of diagnosis. Further details on the model and parameter values are provided in Appendix.

## Data Availability

Processed data and code will be made available in a public repository

## Data availability

Processed data and code will be made available in a public repository upon publication.

## References

1. Koelle K, Martin MA, Antia R, Lopman B, Dean NE. The changing epidemiology of SARS-CoV-2. Science. 2022 Mar 11;375(6585):1116–21.

2. Guzzetta G, Riccardo F, Marziano V, Poletti P, Trentini F, Bella A, et al. Impact of a Nationwide Lockdown on SARS-CoV-2 Transmissibility, Italy. Emerg Infect Dis. 2020/10/20 ed. 2021 Jan;27(1):267–70.

3. Pan A, Liu L, Wang C, Guo H, Hao X, Wang Q, et al. Association of Public Health Interventions With the Epidemiology of the COVID-19 Outbreak in Wuhan, China. JAMA. 2020 May 19;323(19):1915–23.

4. Di Domenico L, Pullano G, Sabbatini CE, Boëlle PY, Colizza V. Impact of lockdown on COVID-19 epidemic in Île-de-France and possible exit strategies. BMC Med. 2020 Jul 30;18(1):240.

5. Coletti P, Libin P, Petrof O, Willem L, Abrams S, Herzog SA, et al. A data-driven metapopulation model for the Belgian COVID-19 epidemic: assessing the impact of lockdown and exit strategies. BMC Infect Dis. 2021 May 30;21(1):503.

6. Marziano V, Guzzetta G, Rondinone BM, Boccuni F, Riccardo F, Bella A, et al. Retrospective analysis of the Italian exit strategy from COVID-19 lockdown. Proc Natl Acad Sci. 2021 Jan 26;118(4):e2019617118.

7. Mathieu E, Ritchie H, Ortiz-Ospina E, Roser M, Hasell J, Appel C, et al. A global database of COVID-19 vaccinations. Nat Hum Behav. 2021 Jul;5(7):947–53.

8. Stefanelli P, Trentini F, Guzzetta G, Marziano V, Mammone A, Schepisi MS, et al. Co-circulation of SARS-CoV-2 Alpha and Gamma variants in Italy, February and March 2021. Eurosurveillance. 2022 Feb 3;27(5):2100429.

9. Davies NG, Abbott S, Barnard RC, Jarvis CI, Kucharski AJ, Munday JD, et al. Estimated transmissibility and impact of SARS-CoV-2 lineage B.1.1.7 in England. Science. 2021/03/03 ed. 2021 Apr 9;372(6538):eabg3055.

10. Alizon S, Haim-Boukobza S, Foulongne V, Verdurme L, Trombert-Paolantoni S, Lecorche E, et al. Rapid spread of the SARS-CoV-2 Delta variant in some French regions, June 2021. Eurosurveillance. 2021 Jul 15;26(28):2100573.

11. Ferguson NM. B.1.617.2 transmission in England: risk factors and transmission advantage. 2021 Jun;14.

12. Fabiani M, Puopolo M, Morciano C, Spuri M, Alegiani SS, Filia A, et al. Effectiveness of mRNA vaccines and waning of protection against SARS-CoV-2 infection and severe covid-19 during predominant circulation of the delta variant in Italy: retrospective cohort study. BMJ. 2022 Feb 10;376:e069052.

13. Andrews N, Stowe J, Kirsebom F, Toffa S, Rickeard T, Gallagher E, et al. Covid-19 Vaccine Effectiveness against the Omicron (B.1.1.529) Variant. N Engl J Med [Internet]. 2022 Mar 2 [cited 2022 Apr 11]; Available from: https://doi.org/10.1056/NEJMoa2119451

14. Fabiani M, Puopolo M, Filia A, Sacco C, Mateo-Urdiales A, Spila Alegiani S, et al. Effectiveness of an mRNA vaccine booster dose against SARS-CoV-2 infection and severe COVID-19 in persons aged ≥60 years and other high-risk groups during predominant circulation of the delta variant in Italy, 19 July to 12 December 2021. Expert Rev Vaccines. 2022 Apr 7;0(0):1–8.

15. Altarawneh HN, Chemaitelly H, Hasan MR, Ayoub HH, Qassim S, AlMukdad S, et al. Protection against the Omicron Variant from Previous SARS-CoV-2 Infection. N Engl J Med. 2022 Mar 31;386(13):1288–90.

16. Hall VJ, Foulkes S, Charlett A, Atti A, Monk EJM, Simmons R, et al. SARS-CoV-2 infection rates of antibody-positive compared with antibody-negative health-care workers in England: a large, multicentre, prospective cohort study (SIREN). The Lancet. 2021 Apr 17;397(10283):1459–69.

17. Lauring AS, Tenforde MW, Chappell JD, Gaglani M, Ginde AA, McNeal T, et al. Clinical severity of, and effectiveness of mRNA vaccines against, covid-19 from omicron, delta, and alpha SARS-CoV-2 variants in the United States: prospective observational study. BMJ. 2022 Mar 9;376:e069761.

18. Sacco C, Mateo-Urdiales A, Petrone D, Spuri M, Fabiani M, Vescio MF, et al. Estimating averted COVID-19 cases, hospitalisations, intensive care unit admissions and deaths by COVID-19 vaccination, Italy, January−September 2021. Eurosurveillance. 2021 Nov 25;26(47):2101001.

19. Bager P, Wohlfahrt J, Bhatt S, Stegger M, Legarth R, Møller CH, et al. Risk of hospitalisation associated with infection with SARS-CoV-2 omicron variant versus delta variant in Denmark: an observational cohort study. Lancet Infect Dis [Internet]. 2022 Apr 22 [cited 2022 Apr 29];0(0). Available from: https://www.thelancet.com/journals/laninf/article/PIIS1473-3099(22)00154-2/fulltext

20. Wolter N, Jassat W, Walaza S, Welch R, Moultrie H, Groome M, et al. Early assessment of the clinical severity of the SARS-CoV-2 omicron variant in South Africa: a data linkage study. The Lancet. 2022 Jan 29;399(10323):437–46.

21. Istituto Superiore di Sanità. Monitoraggio delle varianti del virus SARS-CoV-2 di interesse in sanità pubblica in Italia [Internet]. [cited 2022 Apr 22]. Available from: https://www.epicentro.iss.it/coronavirus/sars-cov-2-monitoraggio-varianti-indagini-rapide

22. Covid-19 Opendata Vaccini [Internet]. Developers Italia; 2022 [cited 2022 Apr 12]. Available from: https://github.com/italia/covid19-opendata-vaccini

23. Task force COVID-19 del Dipartimento Malattie Infettive, Servizio di Informatica, Istituto Superiore di Sanità. Epidemia COVID-19. Aggiornamento nazionale: 2 marzo 2022 [Internet]. Available from: https://www.epicentro.iss.it/coronavirus/bollettino/Bollettino-sorveglianza-integrata-COVID-19_2-marzo-2022.pdf

24. Riccardo F, Ajelli M, Andrianou XD, Bella A, Manso MD, Fabiani M, et al. Epidemiological characteristics of COVID-19 cases and estimates of the reproductive numbers 1 month into the epidemic, Italy, 28 January to 31 March 2020. Eurosurveillance. 2020 Dec 10;25(49):2000790.

25. Istituto Superiore di Sanità. COVID-19 ISS open data – EpiCentro. [Internet]. [cited 2022 Apr 19]. Available from: https://www.epicentro.iss.it/coronavirus/open-data/covid_19-iss.xlsx

26. Poletti P, Tirani M, Cereda D, Trentini F, Guzzetta G, Sabatino G, et al. Association of Age With Likelihood of Developing Symptoms and Critical Disease Among Close Contacts Exposed to Patients With Confirmed SARS-CoV-2 Infection in Italy. JAMA Netw Open. 2021 Mar 10;4(3):e211085.

27. Thomas SJ, Moreira ED, Kitchin N, Absalon J, Gurtman A, Lockhart S, et al. Safety and Efficacy of the BNT162b2 mRNA Covid-19 Vaccine through 6 Months. N Engl J Med. 2021 Nov 4;385(19):1761–73.

28. El Sahly HM, Baden LR, Essink B, Doblecki-Lewis S, Martin JM, Anderson EJ, et al. Efficacy of the mRNA-1273 SARS-CoV-2 Vaccine at Completion of Blinded Phase. N Engl J Med. 2021 Nov 4;385(19):1774–85.

29. Petherick A, Goldszmidt R, Andrade EB, Furst R, Hale T, Pott A, et al. A worldwide assessment of changes in adherence to COVID-19 protective behaviours and hypothesized pandemic fatigue. Nat Hum Behav. 2021 Sep;5(9):1145–60.

30. Campbell F, Archer B, Laurenson-Schafer H, Jinnai Y, Konings F, Batra N, et al. Increased transmissibility and global spread of SARS-CoV-2 variants of concern as at June 2021. Eurosurveillance. 2021 Jun 17;26(24):2100509.

31. Sacco C, Petrone D, Manso MD, Mateo-Urdiales A, Fabiani M, Bressi M, et al. Risk and protective factors for SARS-CoV-2 reinfections, surveillance data, Italy, August 2021 to March 2022. Eurosurveillance. 2022 May 19;27(20):2200372.

32. Salje H, Tran Kiem C, Lefrancq N, Courtejoie N, Bosetti P, Paireau J, et al. Estimating the burden of SARS-CoV-2 in France. Science. 2020 Jul 10;369(6500):208–11.

33. Zardini A, Galli M, Tirani M, Cereda D, Manica M, Trentini F, et al. A quantitative assessment of epidemiological parameters required to investigate COVID-19 burden. Epidemics. 2021 Dec 1;37:100530.

34. Poletti P, Tirani M, Cereda D, Trentini F, Guzzetta G, Marziano V, et al. Age-specific SARS-CoV-2 infection fatality ratio and associated risk factors, Italy, February to April 2020. Eurosurveillance. 2020 Aug 6;25(31):2001383.

35. Khandaker G, Dierig A, Rashid H, King C, Heron L, Booy R. Systematic review of clinical and epidemiological features of the pandemic influenza A (H1N1) 2009. Influenza Other Respir Viruses. 2011 May 1;5(3):148–56.

36. Markov PV, Katzourakis A, Stilianakis NI. Antigenic evolution will lead to new SARS-CoV-2 variants with unpredictable severity. Nat Rev Microbiol. 2022 May;20(5):251–2.

37. Yang J, Marziano V, Deng X, Guzzetta G, Zhang J, Trentini F, et al. Despite vaccination, China needs non-pharmaceutical interventions to prevent widespread outbreaks of COVID-19 in 2021. Nat Hum Behav. 2021 Aug 1;5(8):1009–20.

38. Marziano V, Guzzetta G, Mammone A, Riccardo F, Poletti P, Trentini F, et al. The effect of COVID-19 vaccination in Italy and perspectives for living with the virus. Nat Commun. 2021 Dec 14;12(1):7272.

39. Mossong J, Hens N, Jit M, Beutels P, Auranen K, Mikolajczyk R, et al. Social Contacts and Mixing Patterns Relevant to the Spread of Infectious Diseases. PLOS Med. 2008 Mar 25;5(3):e74.

40. Hu S, Wang W, Wang Y, Litvinova M, Luo K, Ren L, et al. Infectivity, susceptibility, and risk factors associated with SARS-CoV-2 transmission under intensive contact tracing in Hunan, China. Nat Commun. 2021 Mar 9;12(1):1533.

41. Sun K, Wang W, Gao L, Wang Y, Luo K, Ren L, et al. Transmission heterogeneities, kinetics, and controllability of SARS-CoV-2. Science. 2021 Jan 15;371(6526):eabe2424.

42. Diekmann O, Heesterbeek JAP, Metz JAJ. On the definition and the computation of the basic reproduction ratio R0 in models for infectious diseases in heterogeneous populations. J Math Biol. 1990 Jun 1;28(4):365–82.

43. Diekmann O, Heesterbeek J a. P, Roberts MG. The construction of next-generation matrices for compartmental epidemic models. J R Soc Interface. 2010 Jun 6;7(47):873–85.

44. Harris RJ, Hall JA, Zaidi A, Andrews NJ, Dunbar JK, Dabrera G. Effect of Vaccination on Household Transmission of SARS-CoV-2 in England. N Engl J Med. 2021 Aug 19;385(8):759–60.

45. Lipsitch M, Kahn R. Interpreting vaccine efficacy trial results for infection and transmission. Vaccine. 2021 Jul 5;39(30):4082–8.

