## Appendix for "The decline of COVID-19 severity and lethality over two years of pandemic"

### 1. Materials and Methods

#### 1.1 Model for SARS-CoV-2 transmission and vaccination

We developed an age-structured stochastic model for SARS-CoV-2 transmission and vaccination, based on a susceptible-infectious-removed-susceptible scheme (SIRS) and adapted from previously published models (1,2)

The model was used to simulate the evolution of COVID-19 epidemiology in Italy between February 21, 2020, and February 20, 2022.

The simulation period is conventionally subdivided in the following phases:

1. “ancestral (phase 1)”, from February 21 to June 30, 2020. This period was characterized by a first pandemic wave and the circulation of SARS-CoV-2 ancestral lineages. During this phase the Italian health care system was put under substantial pressure (3) and the spread of SARS-CoV-2 was contained through a strict national lockdown (4).
2. “ancestral (phase 2)”, from July 1, 2020, to February 17, 2021. This second phase was characterized by a summer period of low incidence and by a second pandemic wave in the fall of 2020, still caused by ancestral lineages. The second pandemic wave was countered through the introduction of a tier-based reactive restriction system (5). On December 27, 2020, the vaccination campaign was launched.
3. “Alpha (phase 3)”, from February 18, 2021, to July 1, 2021. This phase was characterized by the dominant circulation of the Alpha variant (6) (Table S1) and by a substantial scale-up in COVID-19 vaccination rates (Figure 1b in the main text).
4. “Delta (phase 4)”, from July 2, 2021, to December 23, 2021. This phase was characterized by the dominant circulation of the Delta variant (6) (Table S1) and by the progression of the vaccination campaign, including the administration of booster doses.
5. “Omicron (phase 5)”, from December 24, 2021 to February 20, 2022, characterized by the dominant circulation of the Omicron variant (6) (Table S1).

Mixing patterns were assumed to be heterogeneous across ages according to an age-specific social contact matrix estimated prior to the COVID-19 pandemic (7). We assumed an age-dependent susceptibility to SARS-CoV-2 infection in the ancestral phases: lower in children under 15 years of age and higher for the elderly (65+), compared to individuals of working age (8). For the Alpha, Delta, and Omicron phases susceptibility to SARS-CoV-2 infection was assumed homogeneous across ages.

We simulate a vaccination campaign including the administration of first, second doses and booster doses. Vaccination is assumed to reduce the individuals’ susceptibility to SARS-CoV-2 infection. Breakthrough infections (i.e., infections in vaccinated individuals) are assumed to be half as infectious as those in unvaccinated individuals (9,10).

The model accounts for waning of both protection from natural infection and vaccination. Protection from natural immune response after infection with all lineages is assumed to wane exponentially with a constant rate over the considered period (11). Before waning, natural infection provides complete protection against re-infection with homologous and previous variants, while we assume a partial cross-protection against Omicron infection (12) granted by natural infection with previous variants. Homologous re-infection without waning of natural immunity is not considered for any lineage. In the Delta and Omicron phases we considered waning of protection from vaccine-induced immune response after two doses of vaccine and booster (13–15). Before waning of vaccine protection, vaccination reduces the probability of infection (with different efficacy estimates for the different endpoints and considered strains). After waning of immunity either from natural infection or vaccination, individuals are considered susceptible to infection with the same risk of unvaccinated individuals who were never exposed to SARS-CoV-2. We note that individuals with waned protection against infection may still be protected against other clinical endpoints, such as symptoms, severe disease, or death.

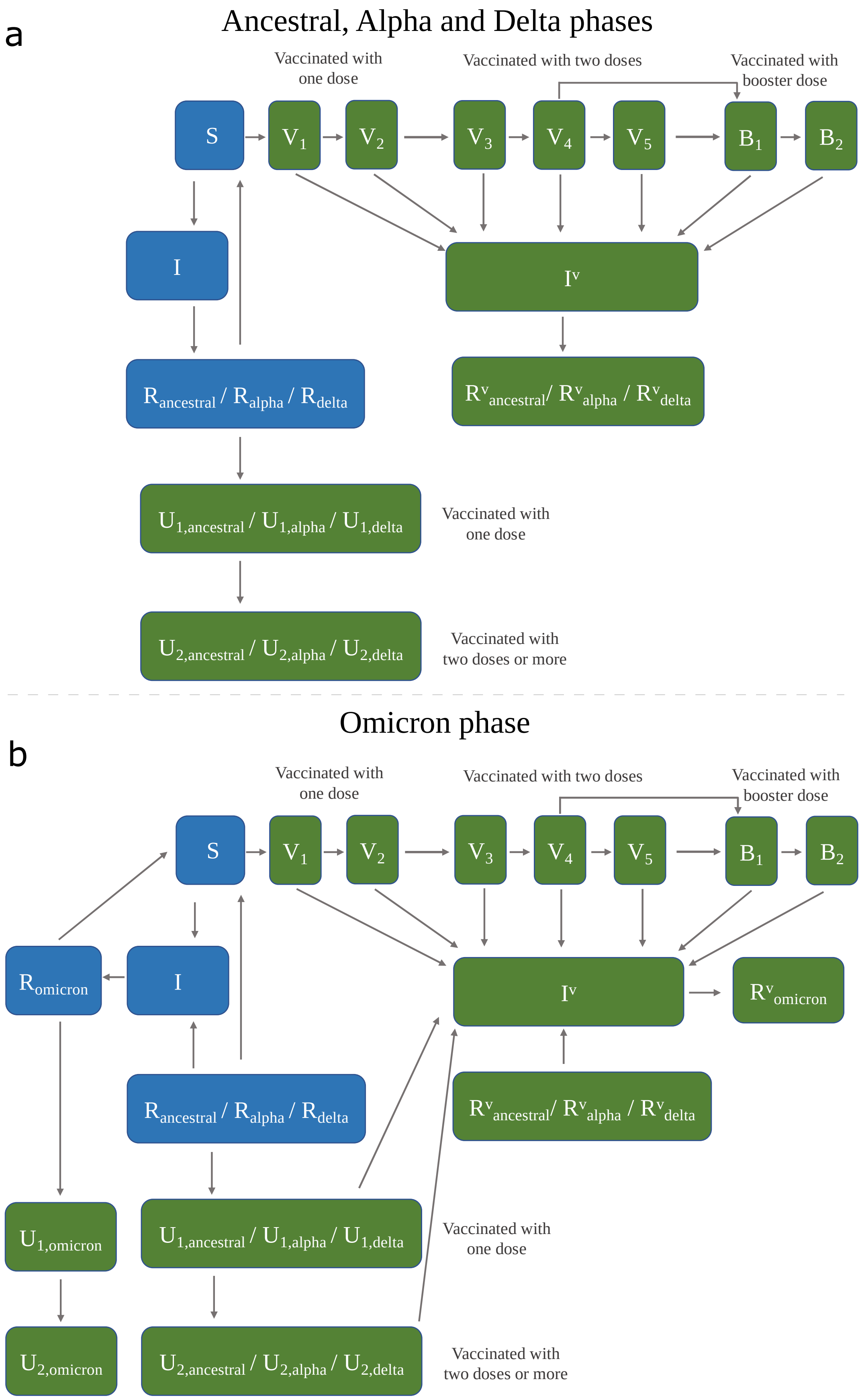

**Figure S1. Schematic representation model transitions.** Blue compartments represent unvaccinated individuals and therefore eligible for vaccination with the first dose; green compartments represent vaccinated individuals. a) ancestral, Alpha and Delta phases. b) Omicron phase.

Model transitions are summarized in the schematic representations in Figure S1 and described in detail by the following system of differential equations

$$
\begin{cases}
S_a'(t) = -\lambda_{S,a}(t)S_a(t) - \alpha_a(t)S_a(t) + \upsilon_R\left[R_{\text{ancestral},a}(t) + R_{\text{alpha},a}(t) + R_{\text{delta},a}(t)\right] \\
I_a'(t) = \lambda_{S,a}(t)S_a(t) - \gamma I_a(t) + f_{\text{omicron}}(t)\lambda_{R,a}(t)[R_{\text{ancestral},a}(t) + R_{\text{alpha},a}(t) + R_{\text{delta},a}(t)] \\
R_{\text{ancestral},a}'(t) = f_{\text{ancestral}}(t)\gamma I_a(t) - \alpha_a(t)R_{\text{ancestral},a}(t) - \upsilon_R R_{\text{ancestral},a}(t) - f_{\text{omicron}}(t)\lambda_{R,a}(t)R_{\text{ancestral},a}(t) \\
R_{\text{alpha},a}'(t) = f_{\text{alpha}}(t)\gamma I_a(t) - \alpha_a(t)R_{\text{alpha},a}(t) - \upsilon_R R_{\text{alpha},a}(t) - f_{\text{omicron}}(t)\lambda_{R,a}(t)R_{\text{alpha},a}(t) \\
R_{\text{delta},a}'(t) = f_{\text{delta}}(t)\gamma I_a(t) - \alpha_a(t)R_{\text{delta},a}(t) - \upsilon_R R_{\text{delta},a}(t) - f_{\text{omicron}}(t)\lambda_{R,a}(t)R_{\text{delta},a}(t) \\
R_{\text{omicron},a}'(t) = f_{\text{omicron}}(t)\gamma I_a(t) - \alpha_a(t)R_{\text{omicron},a}(t) \\
V_{1,a}'(t) = \alpha_a(t)S_a(t) - \lambda_{V_1,a}(t)V_{1,a}(t) - \omega_1 V_{1,a}(t) \\
V_{2,a}'(t) = \omega_1 V_{1,a}(t) - \lambda_{V_2,a}(t)V_{2,a}(t) - \omega_2 V_{2,a}(t) \\
V_{3,a}'(t) = \omega_2 V_{2,a}(t) - \lambda_{V_3,a}(t)V_{3,a}(t) - \omega_3 V_{3,a}(t) \\
V_{4,a}'(t) = \omega_3 V_{3,a}(t) - \lambda_{V_4,a}(t)V_{4,a}(t) - \upsilon_V(t)V_{4,a}(t) - \beta_a(t)V_{4,a}(t) \\
V_{5,a}'(t) = \upsilon_V(t)V_{4,a}(t) - \lambda_{V_5,a}(t)V_{5,a}(t) - \beta_a(t)V_{5,a}(t) \\
B_{1,a}'(t) = \beta_a(t)\left[V_{4,a}(t) + V_{5,a}(t)\right] - \lambda_{B_1,a}(t)B_{1,a}(t) - \upsilon_B(t)B_{1,a}(t) \\
B_{2,a}'(t) = \upsilon_B(t)B_{1,a}(t) - \lambda_{B_2,a}(t)B_{2,a}(t) \\
U_{1,\text{ancestral},a}'(t) = \alpha_a(t)R_{\text{ancestral},a}(t) - [\omega_1\omega_2/(\omega_1+\omega_2)]U_{1,\text{ancestral},a}(t) - f_{\text{omicron}}(t)\lambda_{U_1,a}(t)U_{1,\text{ancestral},a}(t) \\
U_{2,\text{ancestral},a}'(t) = [\omega_1\omega_2/(\omega_1+\omega_2)]U_{1,\text{ancestral},a}(t) - f_{\text{omicron}}(t)\lambda_{U_2,a}(t)U_{2,\text{ancestral},a}(t) \\
U_{1,\text{alpha},a}'(t) = \alpha_a(t)R_{\text{alpha},a}(t) - [\omega_1\omega_2/(\omega_1+\omega_2)]U_{1,\text{alpha},a}(t) - f_{\text{omicron}}(t)\lambda_{U_1,a}(t)U_{1,\text{alpha},a}(t) \\
U_{2,\text{alpha},a}'(t) = [\omega_1\omega_2/(\omega_1+\omega_2)]U_{1,\text{alpha},a}(t) - f_{\text{omicron}}(t)\lambda_{U_2,a}(t)U_{2,\text{alpha},a}(t) \\
U_{1,\text{delta},a}'(t) = \alpha_a(t)R_{\text{delta},a}(t) - [\omega_1\omega_2/(\omega_1+\omega_2)]U_{1,\text{delta},a}(t) - f_{\text{omicron}}(t)\lambda_{U_1,a}(t)U_{1,\text{delta},a}(t) \\
U_{2,\text{delta},a}'(t) = [\omega_1\omega_2/(\omega_1+\omega_2)]U_{1,\text{delta},a}(t) - f_{\text{omicron}}(t)\lambda_{U_2,a}(t)U_{2,\text{delta},a}(t) \\
U_{1,\text{omicron},a}'(t) = \alpha_a(t)R_{\text{omicron},a}(t) - [\omega_1\omega_2/(\omega_1+\omega_2)]U_{1,\text{omicron},a}(t) \\
U_{2,\text{omicron},a}'(t) = [\omega_1\omega_2/(\omega_1+\omega_2)]U_{1,\text{omicron},a}(t) \\
I_a^{V\prime}(t) = \sum_{k=1}^{5}[\lambda_{V_k,a}(t)V_{k,a}(t)] + \sum_{k=1}^{2}[\lambda_{B_k,a}(t)B_{k,a}(t)] + \\
+f_{\text{omicron}}(t)\sum_{k=1}^{2}\left[\lambda_{U_k,a}(t)\left(U_{k,\text{ancestral},a}(t) + U_{k,\text{alpha},a}(t) + U_{k,\text{delta},a}(t)\right)\right] + \\
+f_{\text{omicron}}(t)\lambda_{R^V,a}(t)\left[R_{\text{ancestral},a}^V(t) + R_{\text{alpha},a}^V(t) + R_{\text{delta},a}^V(t)\right] - \gamma I_a^V(t) \\
{R_{\text{ancestral},a}^V}'(t) = f_{\text{ancestral}}(t)\gamma I_a^V(t) - f_{\text{omicron}}(t)\lambda_{R^V,a}(t)R_{\text{ancestral},a}^V(t) \\
{R_{\text{alpha},a}^V}'(t) = f_{\text{alpha}}(t)\gamma I_a^V(t) - f_{\text{omicron}}(t)\lambda_{R^V,a}(t)R_{\text{alpha},a}^V(t) \\
{R_{\text{delta},a}^V}'(t) = f_{\text{delta}}(t)\gamma I_a^V(t) - f_{\text{omicron}}(t)\lambda_{R^V,a}(t)R_{\text{delta},a}^V(t) \\
{R_{\text{omicron},a}^V}'(t) = f_{\text{omicron}}(t)\gamma I_a^V(t)
\end{cases}
$$

where:

- the functions $f_{\text{ancestral}}$; $f_{\text{alpha}}$; $f_{\text{delta}}$ and $f_{\text{omicron}}$ are step functions defined as follows:

$$f_{\text{ancestral}}(t) = \begin{cases} 1, & \mathrm{T}_0 \le t < \mathrm{T}_{\text{alpha}} \\ 0, & \text{otherwise} \end{cases}$$

$$f_{\text{alpha}}(t) = \begin{cases} 1, & \mathrm{T}_{\text{alpha}} \le t < \mathrm{T}_{\text{delta}} \\ 0, & \text{otherwise} \end{cases}$$

$$f_{\text{delta}}(t) = \begin{cases} 1, & \mathrm{T}_{\text{delta}} \le t < \mathrm{T}_{\text{omicron}} \\ 0, & \text{otherwise} \end{cases}$$

$$f_{\text{omicron}}(t) = \begin{cases} 1, & \mathrm{T}_{\text{omicron}} \le t \le \mathrm{T}_{\text{max}} \\ 0, & \text{otherwise} \end{cases}$$

where $\mathrm{T}_0$ is the day of simulation start (February 21, 2020); $\mathrm{T}_{\text{alpha}}$ is the first day of the Alpha phase (February 18, 2021); $\mathrm{T}_{\text{delta}}$ is the first day of the Delta phase (July 2, 2021); $\mathrm{T}_{\text{omicron}}$ is the first day of the Omicron phase (December 24, 2021) and $\mathrm{T}_{\text{max}}$ is the last day of simulations (February 20, 2022). These dates of transition were conventionally defined based on when variants were assumed to become dominant (prevalence >50%), taking reference from genomic surveillance estimates of the prevalence of SARS-CoV-2 lineages (6), reported in in Table S1. The date of transition between the first and the second ancestral phases will be denoted with $\mathrm{T}_{\text{ancestral (phase 2)}}$ and was conventionally defined as July 1st, 2020.

**Table S1.** Prevalence of SARS-CoV-2 variants as estimated from selected flash surveys conducted in Italy (6), with corresponding conventional date of transition to dominance assumed in this study.

| SARS-CoV-2 variant | Last estimate before dominance | | First estimate after dominance | | Date of transition |
|---|---|---|---|---|---|
| | Date | Prevalence | Date | Prevalence | |
| Alpha | Feb 5, 2021 | 17.8% | Feb 18, 2021 | 54.0% | Feb 18, 2021 ($\mathrm{T}_{\text{alpha}}$) |
| Delta | Jun 22, 2021 | 22.7% | Jul 20, 2021 | 94.8% | Jul 2, 2021 ($\mathrm{T}_{\text{delta}}$) |
| Omicron | Dec 20, 2021 | 21.0% | Jan 3, 2022 | 80.8% | Dec 24, 2021 ($\mathrm{T}_{\text{omicron}}$) |

- $S_a(t)$ represents the number of unvaccinated individuals in the age group $a$ who are unprotected against SARS-CoV-2 infection at time $t$.
- $I_a(\mathrm{t})$ represents the number of infectious unvaccinated individuals in the age group $a$ at time $t$. Infectious individuals are assumed to have been infected with ancestral lineages if $t < \mathrm{T}_{\text{alpha}}$, with the Alpha variant if $\mathrm{T}_{\text{alpha}} \leq t < \mathrm{T}_{\text{delta}}$, with Delta if $\mathrm{T}_{\text{delta}} \leq t < \mathrm{T}_{\text{omicron}}$; and with Omicron if $\mathrm{T}_{\text{omicron}} \leq t \leq \mathrm{T}_{\text{max}}$.
- $R_{\text{ancestral},a}(t)$; $R_{\text{alpha},a}(t)$; $R_{\text{delta},a}(t)$; $R_{\text{omicron},a}(t)$ represent the number of unvaccinated individuals in the age group $a$ who at time $t$ have recovered from infection with the different strains and for whom natural immunity has not waned.
- $V_{1,a}(t)$; $V_{2,a}(t)$; $V_{3,a}(t)$; $V_{4,a}(t)$; $V_{5,a}(t)$; $B_{1,a}(t)$ and $B_{2,a}(t)$ represent the number of vaccinated individuals at different stages of protection at time $t$. In particular,
    1) $V_{1,a}$ denotes individuals in the age group $a$ vaccinated with the first dose, for whom the first dose is not effective yet.
    2) $V_{2,a}$ denotes individuals in the age group $a$ vaccinated with the first dose, for whom the first dose is effective.
    3) $V_{3,a}$ denotes individuals in the age group $a$ vaccinated with the second dose for whom the second dose is not effective yet.
    4) $V_{4,a}$ denotes individuals in the age group $a$ vaccinated with the second dose for whom the second dose is effective.
    5) $V_{5,a}$ denotes individuals in the age group $a$ vaccinated with the second dose for whom vaccine protection has waned.
    6) $B_{1,a}$ denotes individuals in the age group $a$ vaccinated with the booster dose for whom the booster dose is effective.
    7) $B_{2,a}$ denotes individuals in the age group $a$ vaccinated with the booster dose for whom vaccine protection has waned.

  Compartments $V_{1,a}$ and $V_{3,a}$encode the delay required for the mounting of an effective immune response.
- $U_{1,\text{ancestral},a}(t)$; $U_{1,\text{alpha},a}(t)$; $U_{1,\text{delta},a}(t)$; $U_{1,\text{omicron},a}(t)$ represent the number of individuals in the age group $a$ who at time $t$ have been vaccinated with the first dose, despite having already experienced SARS-CoV-2 infection with the corresponding strains, and for whom the first dose is effective.
- $U_{2,\text{ancestral},a}(t)$; $U_{2,\text{alpha},a}(t)$; $U_{2,\text{delta},a}(t)$; $U_{2,\text{omicron},a}(t)$ represent the number of individuals in the age group $a$ who at time $t$ are vaccinated with the second dose despite having already experienced SARS-CoV-2 infection with each of the considered strains, and for whom the second dose is effective.
- $I_a^V(t)$ represents the number of infectious individuals in the age group $a$ at time $t$ among those who have already received at least one vaccine dose. Infectious individuals are assumed

to have been infected with ancestral lineages if $t < T_{alpha}$, with the Alpha variant if $T_{alpha} \leq t < T_{delta}$, with Delta if $T_{delta} \leq t < T_{omicron}$ and with Omicron if $T_{omicron} \leq t \leq T_{max}$.

- $R^V_{ancestral,a}(t)$; $R^V_{alpha,a}(t)$; $R^V_{delta,a}(t)$; $R^V_{omicron,a}(t)$ represent the number of individuals in the age group $a$ who at time $t$ have recovered from natural infection with the different strains, contracted after having received at least one vaccine dose.

The model described by the system of ordinary differential equations above is implemented through a stochastic discrete-time model with a time step $\tau = \frac{1}{4} day$.

<u>Force of infection</u>

Unvaccinated individuals who are unprotected against SARS-CoV-2 infection ($S$) are exposed to a time and age-dependent force of infection $\lambda_{S,a}(t)$ which is defined as:

$$\lambda_{S,a}(t) = \delta(t) r_a(t) \sum_{\tilde{a}} C_{a,\tilde{a}} \frac{I_{\tilde{a}}(t) + \pi I^V_{\tilde{a}}(t)}{N_{\tilde{a}}}$$

where:

- $\delta(t)$ is the SARS-CoV-2 transmission rate at time t (see Section 1.2 for details);
- $r_a(t)$ is the relative susceptibility to SARS-CoV-2 infection at age $a$. For ancestral phases ($t < T_{alpha}$), we sample susceptibility profiles from the posterior distribution estimated in (8), having mean values $r_a(t)$=0.58 (95%CI 0.34-0.98) under 15 years of age; $r_a(t)$=1 between 15 and 64 years; and $r_a(t)$=1.65 (95%CI 1.03-2.65) above 64 years. From the Alpha phase on ($t \geq T_{alpha}$), we assume $r_a(t)$=1 for all ages.
- $C_{a,\tilde{a}}$ represents the age-group-specific contact matrix, as estimated before the SARS-CoV-2 pandemic (7), whose entries describe the mean numbers of persons in age group $\tilde{a}$ encountered by an individual of age group $a$ in an average day.
- $\pi$ represents the relative infectiousness of SARS-CoV-2 infections among vaccinated compared to unvaccinated. We assumed $\pi = 0.5$ (9,10).
- $N_{\tilde{a}}$ represents the number of individuals in the age group $\tilde{a}$.

Vaccinated individuals can get infected with any SARS-CoV-2 variant with a susceptibility that depends on vaccination stage and the effectiveness of the vaccine against the circulating variant. Individuals who have recovered from natural infection with ancestral lineages, Alpha or Delta variant may get re-infected with the Omicron variant due to its ability to escape immunity from natural infection with heterologous lineages (12).

We modeled the age-dependent force of infection $\lambda_{D,a}(t)$ for individuals in compartment $D$ as follows:

$$\lambda_{D,a}(t) = (1 - \chi_D(t))\, \lambda_{S,a}(t)$$

where $\chi_D$(t) is a time-dependent scaling factor accounting for the reduction in susceptibility of compartment $D$ compared to unprotected unvaccinated individuals ($S$) at time $t$. For vaccinated compartments, the scaling factor $\chi_D$(t) represents the effectiveness of vaccines in preventing infection; for unvaccinated compartments recovered from natural infection with ancestral lineages, Alpha or Delta variants ($R_{ancestral}$; $R_{alpha}$ and $R_{delta}$), $\chi_D$(t) represents the effectiveness of infection with previous lineages in preventing reinfection with Omicron. Values assumed for $\chi_D(t)$ are reported in Table S2.

Protection from natural immune response in unvaccinated individuals recovered from SARS-CoV-2 infection is assumed to wane exponentially with a constant rate $\upsilon_R$=1/730 days$^{-1}$ (11). For all infectious compartments and different phases, the average duration of infectiousness $(1/\gamma)$ is set equal to the average generation time (6.6 days) (16–18).

#### Allocation of vaccine doses

The rollout of the two-dose and booster vaccination campaigns is modeled using detailed data on the daily age-specific number of first and booster doses administered over the considered period (19). The priority order of the Italian vaccination campaign was based on the WHO SAGE roadmap (20) prioritizing high-risk population age segments, i.e. over 80 years of age and essential workers (e.g. health care workers and teachers) and then progressively targeting younger age groups. The same priority order was also followed during rollout of the booster vaccination campaign, started in autumn 2021.

In the model, we assume that the first dose of vaccination is administered to unvaccinated individuals who are unprotected against SARS-CoV-2 infection or have recovered from natural infection with all considered strains.

At each time $t$, the number of unvaccinated individuals in the age group $a$ who will receive a first vaccine dose is determined as a fraction $z_a(t)$ of the corresponding population:

$$z_a(t) = \frac{d_a(t)}{S_a(t) + R_a(t)}$$

where $d_a(t)$ represents the number of first vaccine doses administered to individuals in the age group $a$ at time $t$; and $R_a(t) = R_{\text{ancestral},a} + R_{\text{alpha},a} + R_{\text{delta},a} + R_{\text{omicron},a}$. The value of $d_a(t)$ is inferred from data on the daily number of first doses administered by age group in Italy between the start of vaccination (December 27, 2020) and the end of the simulated period (February 20, 2022) (19).
The first-dose vaccination rate $\alpha_a(t)$ associated to the probability $z_a(t)$ in the differential equations model can be computed trough the following equation:

$$z_a(t) = 1 - e^{-\alpha_a(t)\tau}.$$

We assume that the first dose becomes effective on average after $1/\omega_1 = 14$ days from its administration (15,21). The second dose is administered to individuals who were vaccinated with one dose (and did not contracted infection in the meanwhile) on average 28 days after the first dose (i.e. $1/\omega_1 + 1/\omega_2 = \frac{\omega_1+\omega_2}{\omega_1\omega_2} = 28$ days and $1/\omega_2 = 14$ days) (22). We assume that the second dose becomes effective after $1/\omega_3 = 14$ days from its administration (15,21).

Booster doses are administered to individuals who have received two doses of vaccination. At each time $t$, the number of individuals in the age group $a$ who will receive a booster vaccine dose is determined as a fraction $w_a(t)$ of the corresponding population:

$$w_a(t) = \frac{b_a(t)}{V_{4,a}(t) + V_{5,a}(t) + U_{2,a}(t) + R_a^V(t)}$$

where $b_a(t)$ represents the number of booster doses administered to individuals in the age group $a$ at time $t$; $U_{2,a}(t) = U_{2,\text{ancestral},a}(t) + U_{2,\text{alpha},a}(t) + U_{2,\text{delta},a}(t) + U_{2,\text{omicron},a}(t)$ and $R_a^V(t) = R_{\text{ancestral},a}^V(t) + R_{\text{alpha},a}^V(t) + R_{\text{delta},a}^V(t) + R_{\text{omicron},a}^V(t)$. The value of $b_a(t)$ is inferred from data on the daily number of booster doses administered by age group in Italy in the simulation period (19).

The booster-dose vaccination rate $\beta_a(t)$ associated to the probability $w_a(t)$ in the differential equations model can be computed through the following equation:

$$w_a(t) = 1 - e^{-\beta_a(t)\tau}.$$

First-dose vaccination and booster vaccination coverage by age over the simulated period are shown in Figure S2.

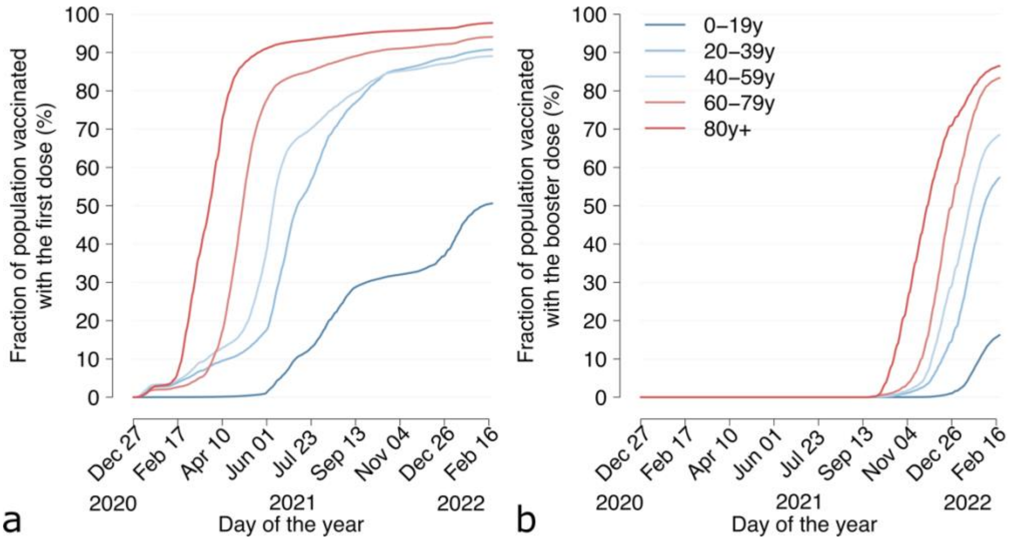

**Figure S2. Vaccination coverage by age group as observed in Italy between December 27, 2020, and February 20, 2022 (19).** a) first dose; b) booster dose. In Italy, administration of two doses is recommended to all individuals aged 5 years or more; administration of a booster dose is recommended to all individuals aged 12 years or more.

We consider waning of protection after two doses of vaccine and after the booster dose (13–15,18). Average durations of protection after two doses and after one booster dose considered for different variants $p$ are reported in Table S2. Due to the different waning rates of vaccine protection across variants, the proportion of vaccine-protected individuals changes when a new variant becomes dominant. For example, the proportion of vaccinees with booster of age $a$ that were protected against Omicron infection at the time of its dominance $T_{\text{omicron}}$ is approximated by

$$P_a = \int_0^{T_{\text{omicron}}} b_a(t) e^{-(T_{\text{omicron}}-t)\upsilon_{B,\text{omicron}}} dt.$$

Since there were

$$Q_a = \int_0^{T_{\text{omicron}}-1} b_a(t) e^{-(T_{\text{omicron}}-1-t)\upsilon_{B,\text{delta}}} dt$$

individuals of age $a$ that were protected against Delta infection at date $T_{\text{omicron}} - 1$, on day $T_{\text{omicron}}$ we move $Q_a - P_a$ individuals from compartment $B_{1,a}$ (booster protected) to compartment $B_{2,a}$ (booster waned). We perform the same operation for all variants and for the two-dose compartment as well.

**Table S2.** Description of key parameters and assumptions used in the model. If not specified, values are set equal to those reported in the column "ancestral phases".

| Parameter | Ancestral, phases 1 and 2 ($t < T_{alpha}$) | Alpha, phase 3 ($T_{alpha} \leq t < T_{delta}$) | Delta, phase 4 ($T_{delta} \leq t < T_{omicron}$) | Omicron, phase 5 ($t \geq T_{omicron}$) | Source |
|---|---|---|---|---|---|
| **Epidemiological** | | | | | |
| Generation time ($1/\gamma$) | 6.6 days | - | - | - | (16) |
| Susceptibility to infection ($r_a(t)$) | Age-dependent<br>- $r_a(t)$=0.58 (95%CI 0.34-0.98) under 15 years;<br>- $r_a(t)$=1 between 15 and 64 years;<br>- $r_a(t)$=1.65 (95%CI 1.03-2.65) over 64 years | Homogenous:<br>$r_a(t)$=1 for all ages | Homogenous:<br>$r_a(t)$=1 for all ages | Homogenous:<br>$r_a(t)$=1 for all ages | (8) |
| Average duration of immunity after natural infection ($1/\upsilon_R$) | 2 years | 2 years | 2 years | - | (11) |
| Age-group specific contact matrix ($C_{a,\tilde{a}}$) | Contact matrix estimated for Italy before the pandemic | - | - | - | (7) |
| **Vaccination** | | | | | |
| Delay between 1st dose and achievement of vaccine efficacy ($1/\omega_1$) | 14 days | - | - | - | (13) |
| Interval between 1st and 2nd dose ($1/\omega_1 + 1/\omega_2$) | 28 days | - | - | - | (22) |
| Delay between 2nd dose and achievement of vaccine efficacy ($1/\omega_3$) | 14 days | - | - | - | (13) |
| Duration of vaccine protection after two doses of vaccine ($1/\upsilon_V(t)$) | no waning observed ($\upsilon_V(t) = \upsilon_{V,\text{ancestral}} = 0$) | no waning observed ($\upsilon_V(t) = \upsilon_{V,\text{alpha}} = 0$) | $1/\upsilon_{V,\text{delta}}$ =200.6 days | $1/\upsilon_{V,\text{omicron}}$ =74.5 days | (18) |
| Duration of vaccine protection after booster ($1/\upsilon_B(t)$) | no waning observed ($\upsilon_B(t) = \upsilon_{B,\text{ancestral}} = 0$) | no waning observed ($\upsilon_B(t) = \upsilon_{B,\text{alpha}} = 0$) | no waning observed ($\upsilon_B(t) = \upsilon_{B,\text{delta}} = 0$) | $1/\upsilon_{B,\text{omicron}}$ = 195.3 days | (18) |
| Relative infectiousness of SARS-CoV-2 breakthrough infections ($\pi$) | 50% | - | - | - | (9,10) |
| **Susceptibility reduction in compartment D compared to unvaccinated individuals who are unprotected against SARS-CoV-2 ($\chi_D(t)$)** | | | | | |
| $\chi_{V_1}(t) = \chi_{V_5}(t) = \chi_{B_2}(t)$ | 0 | 0 | 0 | 0 | Assumed |
| $\chi_{V_2}(t) = \chi_{V_3}(t)$ | 0.49* | 0.49 | 0.502 | 0.428 | (13,14) |
| $\chi_{V_4}(t)$ | 0.79* | 0.79 | 0.69 | 0.655 | (13,14) |
| $\chi_{B_1}(t)$ | 0 | 0 | 0.761 | 0.669 | (14,15) |
| $\chi_{U_{1,\text{ancestral}}}(t) = \chi_{U_{1,\text{alpha}}}(t) = \chi_{U_{1,\text{delta}}}(t) = \chi_{U_{2,\text{ancestral}}}(t) = \chi_{U_{2,\text{alpha}}}(t) = \chi_{U_{2,\text{delta}}}(t) = \chi_{R^V_{\text{ancestral}}}(t) = \chi_{R^V_{\text{alpha}}}(t) = \chi_{R^V_{\text{delta}}}(t)$ | 1 | 1 | 1 | 0.669** | Assumed |
| $\chi_{R_{\text{ancestral}}}(t) = \chi_{R_{\text{alpha}}}(t) = \chi_{R_{\text{delta}}}(t)$ | 1 | 1 | 1 | 0.56 | (12) |

*assumed equal to estimates available for the Alpha phase **assumed equal to estimates of protection from booster in the Omicron phase

#### 1.2 Reproducing the COVID-19 epidemic trajectory in Italy

The reproduction number associated to the dynamical system above can be computed as the dominant eigenvalue of the Next Generation Matrix (NGM) (23–25), defined as:

$$NGM = \frac{\delta(t)}{\gamma}\begin{pmatrix}
B^{S,i_S}_{a,\tilde{a}} & B^{S,i_{V_1}}_{a,\tilde{a}} & B^{S,i_{V_2}}_{a,\tilde{a}} & B^{S,i_{V_3}}_{a,\tilde{a}} & B^{S,i_{V_4}}_{a,\tilde{a}} & B^{S,i_{V_5}}_{a,\tilde{a}} & B^{S,i_{B_1}}_{a,\tilde{a}} & B^{S,i_{B_2}}_{a,\tilde{a}} & B^{S,i_{U_1}}_{a,\tilde{a}} & B^{S,i_{U_2}}_{a,\tilde{a}} & B^{S,i_{R_V}}_{a,\tilde{a}} & B^{S,i_R}_{a,\tilde{a}} \\
B^{V_1,i_S}_{a,\tilde{a}} & B^{V_1,i_{V_1}}_{a,\tilde{a}} & B^{V_1,i_{V_2}}_{a,\tilde{a}} & B^{V_1,i_{V_3}}_{a,\tilde{a}} & B^{V_1,i_{V_4}}_{a,\tilde{a}} & B^{V_1,i_{V_5}}_{a,\tilde{a}} & B^{V_1,i_{B_1}}_{a,\tilde{a}} & B^{V_1,i_{B_2}}_{a,\tilde{a}} & B^{V_1,i_{U_1}}_{a,\tilde{a}} & B^{V_1,i_{U_2}}_{a,\tilde{a}} & B^{V_1,i_{R_V}}_{a,\tilde{a}} & B^{V_1,i_R}_{a,\tilde{a}} \\
B^{V_2,i_S}_{a,\tilde{a}} & B^{V_2,i_{V_1}}_{a,\tilde{a}} & B^{V_2,i_{V_2}}_{a,\tilde{a}} & B^{V_2,i_{V_3}}_{a,\tilde{a}} & B^{V_2,i_{V_4}}_{a,\tilde{a}} & B^{V_2,i_{V_5}}_{a,\tilde{a}} & B^{V_2 i_{B_1}}_{a,\tilde{a}} & B^{V_2,i_{B_2}}_{a,\tilde{a}} & B^{V_2,i_{U_1}}_{a,\tilde{a}} & B^{V_2,i_{U_2}}_{a,\tilde{a}} & B^{V_2,i_{R_V}}_{a,\tilde{a}} & B^{V_2,i_R}_{a,\tilde{a}} \\
B^{V_3,i_S}_{a,\tilde{a}} & B^{V_3,i_{V_1}}_{a,\tilde{a}} & B^{V_3,i_{V_2}}_{a,\tilde{a}} & B^{V_3,i_{V_3}}_{a,\tilde{a}} & B^{V_3,i_{V_4}}_{a,\tilde{a}} & B^{V_3,i_{V_5}}_{a,\tilde{a}} & B^{V_3,i_{B_1}}_{a,\tilde{a}} & B^{V_3,i_{B_2}}_{a,\tilde{a}} & B^{V_3,i_{U_1}}_{a,\tilde{a}} & B^{V_3,i_{U_2}}_{a,\tilde{a}} & B^{V_3,i_{R_V}}_{a,\tilde{a}} & B^{V_3,i_R}_{a,\tilde{a}} \\
B^{V_4,i_S}_{a,\tilde{a}} & B^{V_4,i_{V_1}}_{a,\tilde{a}} & B^{V_4,i_{V_2}}_{a,\tilde{a}} & B^{V_4,i_{V_3}}_{a,\tilde{a}} & B^{V_4,i_{V_4}}_{a,\tilde{a}} & B^{V_4,i_{V_5}}_{a,\tilde{a}} & B^{V_4,i_{B_1}}_{a,\tilde{a}} & B^{V_4,i_{B_2}}_{a,\tilde{a}} & B^{V_4,i_{U_1}}_{a,\tilde{a}} & B^{V_4,i_{U_2}}_{a,\tilde{a}} & B^{V_4,i_{R_V}}_{a,\tilde{a}} & B^{V_4,i_R}_{a,\tilde{a}} \\
B^{V_5,i_S}_{a,\tilde{a}} & B^{V_5,i_{V_1}}_{a,\tilde{a}} & B^{V_5,i_{V_2}}_{a,\tilde{a}} & B^{V_5,i_{V_3}}_{a,\tilde{a}} & B^{V_5,i_{V_4}}_{a,\tilde{a}} & B^{V_5,i_{V_5}}_{a,\tilde{a}} & B^{V_5,i_{B_1}}_{a,\tilde{a}} & B^{V_5,i_{B_2}}_{a,\tilde{a}} & B^{V_5,i_{U_1}}_{a,\tilde{a}} & B^{V_5,i_{U_2}}_{a,\tilde{a}} & B^{V_5,i_{R_V}}_{a,\tilde{a}} & B^{V_5,i_R}_{a,\tilde{a}} \\
B^{B_1,i_S}_{a,\tilde{a}} & B^{B_1,i_{V_1}}_{a,\tilde{a}} & B^{B_1,i_{V_2}}_{a,\tilde{a}} & B^{B_1,i_{V_3}}_{a,\tilde{a}} & B^{B_1,i_{V_4}}_{a,\tilde{a}} & B^{B_1,i_{V_5}}_{a,\tilde{a}} & B^{B_1,i_{B_1}}_{a,\tilde{a}} & B^{B_1,i_{B_2}}_{a,\tilde{a}} & B^{B_1,i_{U_1}}_{a,\tilde{a}} & B^{B_1,i_{U_2}}_{a,\tilde{a}} & B^{B_1,i_{R_V}}_{a,\tilde{a}} & B^{B_1,i_R}_{a,\tilde{a}} \\
B^{B_2,i_S}_{a,\tilde{a}} & B^{B_2,i_{V_1}}_{a,\tilde{a}} & B^{B_2,i_{V_2}}_{a,\tilde{a}} & B^{B_2,i_{V_3}}_{a,\tilde{a}} & B^{B_2,i_{V_4}}_{a,\tilde{a}} & B^{B_2,i_{V_5}}_{a,\tilde{a}} & B^{B_2,i_{B_1}}_{a,\tilde{a}} & B^{B_2,i_{B_2}}_{a,\tilde{a}} & B^{B_2,i_{U_1}}_{a,\tilde{a}} & B^{B_2,i_{U_2}}_{a,\tilde{a}} & B^{B_2,i_{R_V}}_{a,\tilde{a}} & B^{B_2,i_R}_{a,\tilde{a}} \\
B^{U_1,i_S}_{a,\tilde{a}} & B^{U_1,i_{V_1}}_{a,\tilde{a}} & B^{U_1,i_{V_2}}_{a,\tilde{a}} & B^{U_1,i_{V_3}}_{a,\tilde{a}} & B^{U_1,i_{V_4}}_{a,\tilde{a}} & B^{U_1,i_{V_5}}_{a,\tilde{a}} & B^{U_1,i_{B_1}}_{a,\tilde{a}} & B^{U_1,i_{B_2}}_{a,\tilde{a}} & B^{U_1,i_{U_1}}_{a,\tilde{a}} & B^{U_1,i_{U_2}}_{a,\tilde{a}} & B^{U_1,i_{R_V}}_{a,\tilde{a}} & B^{U_1,i_R}_{a,\tilde{a}} \\
B^{U_2,i_S}_{a,\tilde{a}} & B^{U_2,i_{V_1}}_{a,\tilde{a}} & B^{U_2,i_{V_2}}_{a,\tilde{a}} & B^{U_2,i_{V_3}}_{a,\tilde{a}} & B^{U_2,i_{V_4}}_{a,\tilde{a}} & B^{U_2,i_{V_5}}_{a,\tilde{a}} & B^{U_2,i_{B_1}}_{a,\tilde{a}} & B^{U_2,i_{B_2}}_{a,\tilde{a}} & B^{U_2,i_{U_1}}_{a,\tilde{a}} & B^{U_2,i_{U_2}}_{a,\tilde{a}} & B^{U_2,i_{R_V}}_{a,\tilde{a}} & B^{U_2,i_R}_{a,\tilde{a}} \\
B^{R_V,i_S}_{a,\tilde{a}} & B^{U_2,i_{V_1}}_{a,\tilde{a}} & B^{R_V,i_{V_2}}_{a,\tilde{a}} & B^{R_V,i_{V_3}}_{a,\tilde{a}} & B^{R_V,i_{V_4}}_{a,\tilde{a}} & B^{R_V,i_{V_5}}_{a,\tilde{a}} & B^{R_V,i_{B_1}}_{a,\tilde{a}} & B^{R_V,i_{B_2}}_{a,\tilde{a}} & B^{R_V,i_{U_1}}_{a,\tilde{a}} & B^{R_V,i_{U_2}}_{a,\tilde{a}} & B^{R_V,i_{R_V}}_{a,\tilde{a}} & B^{R_V,i_R}_{a,\tilde{a}} \\
B^{R,i_S}_{a,\tilde{a}} & B^{R,i_{V_1}}_{a,\tilde{a}} & B^{R,i_{V_2}}_{a,\tilde{a}} & B^{R,i_{V_3}}_{a,\tilde{a}} & B^{R,i_{V_4}}_{a,\tilde{a}} & B^{R,i_{V_5}}_{a,\tilde{a}} & B^{R,i_{B_1}}_{a,\tilde{a}} & B^{R,i_{B_2}}_{a,\tilde{a}} & B^{R,i_{U_1}}_{a,\tilde{a}} & B^{R,i_{U_2}}_{a,\tilde{a}} & B^{R,i_{R_V}}_{a,\tilde{a}} & B^{R,i_R}_{a,\tilde{a}}
\end{pmatrix} \quad (1)$$

Each block $B^{D,i_D}_{a,\tilde{a}}$ describes the time-dependent contribution to the transmission of age-specific interactions between susceptible individuals in compartment $D$ and infectious individuals who were infected while being in compartment $D$ (here denoted as $i_D$).
Specifically, the explicit computation of the NGM starting from model equations yields:

$$B^{D,i_D}_{a,\tilde{a}}(t) = r_a(t) C_{a,\tilde{a}} [1 - \chi_D(t)]\, \Theta_{i_D} \frac{N^D_{\tilde{a}}(t)}{N_{\tilde{a}}}$$

where:

- $r_a(t)$ is the relative susceptibility to SARS-CoV-2 infection at age $a$;
- $C_{a,\tilde{a}}$ is the age-group-specific contact matrix (7),
- $\chi_D(t)$ is the reduction in susceptibility to SARS-CoV-2 of compartment $D$ compared to unprotected unvaccinated individuals $S$ and $\chi_S(t)$ is set to 0 by definition;
- $\Theta_{i_D}$ is the relative infectiousness of SARS-CoV-2 infections among vaccinated compared to unvaccinated (equal to $\pi$ for vaccinated compartments, and 1 for unvaccinated compartments);
- $N^D_{\tilde{a}}(t)$ is the number of individuals of age $\tilde{a}$ in compartment $D$ at time $t$;
- $N_{\tilde{a}}$ represents the total population of age $\tilde{a}$.

To reproduce the epidemic trajectory observed in Italy over the study period we use Equation 1 to recalibrate daily the value of $\delta(t)$, given the distribution of the susceptibility profile by age ($r_a(t)$), the distribution of the bootstrapped contact matrix ($C_{a,\tilde{a}}$), and the values of $\gamma$, $\chi_D(t)$ and $\pi$ (see Table S2). For each day $t$, the selected value of $\delta(t)$ will be the one that will make the model's reproduction number (recomputed after updating the $N^D_{\tilde{a}}(t)$ to current state variables of the model) match the corresponding value of the net reproduction number as estimated on the same day from epidemic curves collected by the national integrated surveillance system (26,27). Results discussed in the main text and in the following sections were obtained by running 300 simulations, sampling at each run a different value from the joint distribution of the bootstrapped contact matrices $C_{a,\tilde{a}}$ and the relative susceptibility by age $r_a(t)$.

#### 1.3 Model outputs

The main model outcomes are the age-specific number of new infections per day $y_a^D(\mathrm{t})$ among individuals of age group $a$ in compartment $D$ and the state variables (defining the SARS-CoV-2 susceptibility profile of the Italian population) at the end of each considered phase. For each model outcome, we report the mean values and 95% confidence intervals across stochastic simulations.

<u>SARS-CoV-2 infection attack rate and infection ascertainment ratio</u>

We computed the cumulative number of SARS-CoV-2 infections in the age group $a$ in the different phases as:

$$y_{a,\text{ancestral (phase 1)}} = \sum_{t=T_0}^{T_{\text{ancestral (phase 2)}}-1} \sum_D y_a^D(\mathrm{t})$$

$$y_{a,\text{ancestral (phase 2)}} = \sum_{t=T_{\text{ancestral (phase 2)}}}^{T_{\text{alpha}}-1} \sum_D y_a^D(\mathrm{t})$$

$$y_{a,\text{alpha}} = \sum_{t=T_{\text{alpha}}}^{T_{\text{delta}}-1} \sum_D y_a^D(\mathrm{t})$$

$$y_{a,\text{delta}} = \sum_{t=T_{\text{delta}}}^{T_{\text{omicron}}-1} \sum_D y_a^D(\mathrm{t})$$

$$y_{a,\text{omicron}} = \sum_{t=T_{\text{omicron}}}^{T_{\text{max}}} \sum_D y_a^D(\mathrm{t})$$

The age-specific SARS-CoV-2 infection attack rate in the phase $p$ was then computed as $\mathrm{AR}_{a,p} = y_{a,p}/\mathrm{pop}_a$, where $\mathrm{pop}_a$ is the Italian population in age group $a$ (28).

The overall attack rate in phase $p$ is computed as the ratio between the total number of SARS-CoV-2 infections estimated by the model in the same phase, $\mathrm{Y}_p$, and the total Italian population:

$$\mathrm{AR}_{tot,p} = \frac{\mathrm{Y_p}}{\sum_a \mathrm{pop}_a} = \frac{\sum_a y_{a,p}}{\sum_a \mathrm{pop}_a}$$

The infection ascertainment ratio $\rho_p$ in phase $p$ was computed as the ratio between the number of infections reported to the national integrated surveillance system with a date of diagnosis in phase $p$ ($x_{\mathrm{INF},p}$ in Table S3) and the total number of SARS-CoV-2 infections estimated by the model in the same phase, $\mathrm{Y}_p$.

<u>Probabilities of hospitalization, admission to intensive care units (ICU) and death.</u>

For each phase $p$, we computed the probability of hospitalization ($\mathrm{L}_{\mathrm{HOSP},p}$), ICU admission ($\mathrm{L}_{\mathrm{ICU},p}$) and death ($\mathrm{L}_{\mathrm{DEATH},p}$) after SARS-CoV-2 infection as follows:

$$\mathrm{L}_{\mathrm{HOSP},p} = \frac{x_{\mathrm{HOSP},p}}{\mathrm{Y}_p} \qquad \mathrm{L}_{\mathrm{ICU},p} = \frac{x_{\mathrm{ICU},p}}{\mathrm{Y}_p} \qquad \mathrm{L}_{\mathrm{DEATH},p} = \frac{x_{\mathrm{DEATH},p}}{\mathrm{Y}_p}$$

where $x_{\mathrm{HOSP},p}$, $x_{\mathrm{ICU},p}$ and $x_{\mathrm{DEATH},p}$ represent the number of SARS-CoV-2 infections reported to the national integrated surveillance system during phase $p$ that were admitted to the hospital, to the ICU, or died, respectively (Table S3).

**Table S3.** Number of SARS-CoV-2 confirmed infections as reported to the national integrated surveillance system during phase $p$ and those who were admitted to the hospital, to the ICU, or died, respectively, during phase $p$ (26,29).

| Phase ($p$) | Confirmed infections ($x_{INF,p}$) | Hospitalized ($x_{\mathrm{HOSP},p}$) | Admitted to ICU ($x_{\mathrm{HOSP},p}$) | Deaths ($x_{\mathrm{DEATH},p}$) |
|---|---|---|---|---|
| Ancestral (phase 1) | 240,928 | 87,351 | 10,421 | 35,946 |
| Ancestral (phase 2) | 2,561,524 | 207,772 | 29,709 | 66,177 |
| Alpha (phase 3) | 1,467,707 | 117,649 | 16,789 | 25,758 |
| Delta (phase 4) | 1,359,605 | 56,246 | 5,931 | 10,390 |
| Omicron (phase 5) | 6,822,724 | 78,469 | 5,187 | 16,149 |

Relative reductions in the probability of the different outcomes compared to the first ancestral phase were computed as follows:

$$\mathrm{RR}_{\mathrm{HOSP},p} = \frac{\mathrm{L}_{\mathrm{HOSP},p} - \mathrm{L}_{\mathrm{HOSP,ancestral\ (phase\ 1)}}}{\mathrm{L}_{\mathrm{HOSP,ancestral\ (phase\ 1)}}}$$

$$\mathrm{RR}_{\mathrm{ICU},p} = \frac{\mathrm{L}_{\mathrm{ICU},p} - \mathrm{L}_{\mathrm{ICU,ancestral\ (phase\ 1)}}}{\mathrm{L}_{\mathrm{ICU,ancestral\ (phase\ 1)}}}$$

$$\mathrm{RR}_{\mathrm{DEATH},p} = \frac{\mathrm{L}_{\mathrm{DEATH},p} - \mathrm{L}_{\mathrm{DEATH,ancestral\ (phase\ 1)}}}{\mathrm{L}_{\mathrm{DEATH,ancestral\ (phase\ 1)}}}$$

#### 1.4 Model initialization

The population of the model was initialized according to estimates of the Italian population by age at the end of 2020 (28) and approximated as constant throughout the two years of simulations. At simulation start, SARS-CoV-2 infection is seeded in a fully susceptible population and the number of initially infectious individuals is determined in such a way to match COVID-19 deaths reported by the surveillance system in the first ancestral phase (Table S3 and Figure S3). Specifically, the age-specific number of COVID-19 deaths associated to the model in the first ancestral phase is computed as:

$$M_{a,\mathrm{ancestral\ (phase\ 1)}} = \mu_a y_{a,\mathrm{ancestral\ (phase\ 1)}}$$

where $\mu_a$ is the age-specific infection fatality ratio estimated for Italy in the first pandemic wave (30).

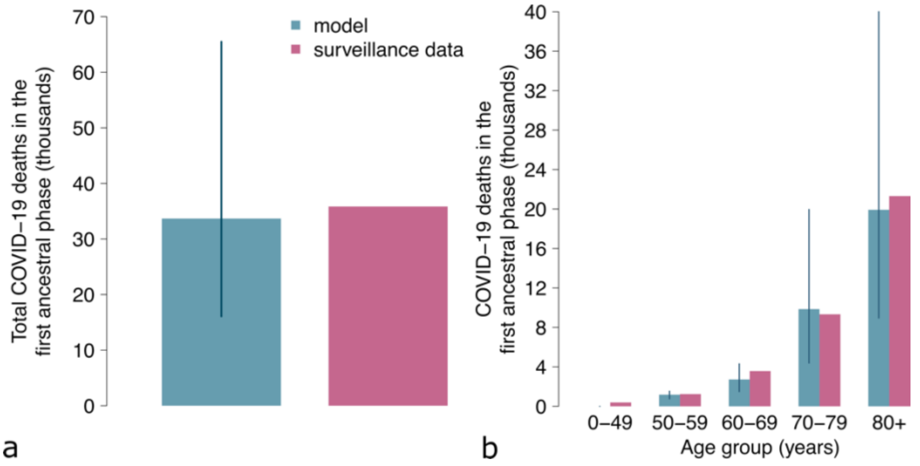

**Figure S3. COVID-19 deaths over the first ancestral phase (in thousands).** Blue: mean (bar) and 95% CI (vertical lines) of the model estimates (n=300 stochastic model realizations); red: data from the Italian Integrated Surveillance System (26,29). **a)** Total; **b)** by age group.

#### 2. Additional results

We compared the age distribution of SARS-CoV-2 confirmed infections reported to the Italian National Integrated Surveillance system in the different phases considered to that of SARS-CoV-2 infections estimated by the model (Figure S4). Both the data and model estimates suggest a gradual shift of infections towards younger age groups.

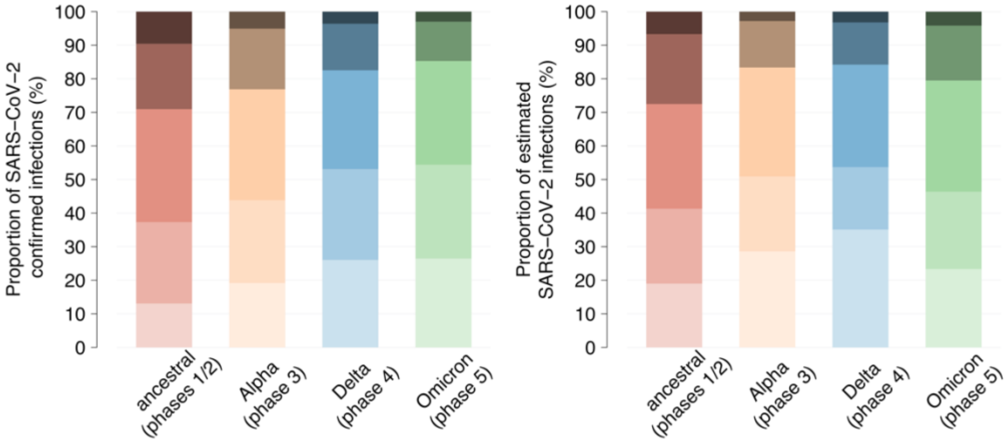

**Figure S4. a)** Age distribution of SARS-CoV-2 confirmed infections reported to the national integrated surveillance system in the ancestral phases (red), in the Alpha phase (orange), in the Delta phase (blue) and in the Omicron phase (green) (29). **b)** Mean age distribution of SARS-CoV-2 infections as estimated by the model in the different phases.

Figure S5 shows the number of COVID-19 tests (per 1,000 individuals) administered in Italy over time, to assist interpretation of the changing ascertainment ratio over time found by our model. The testing generally increased throughout the pandemic and reached peak values during the Omicron phase, where testing capacity was scaled up by over 3 times compared to a previous peak during the Alpha phase.

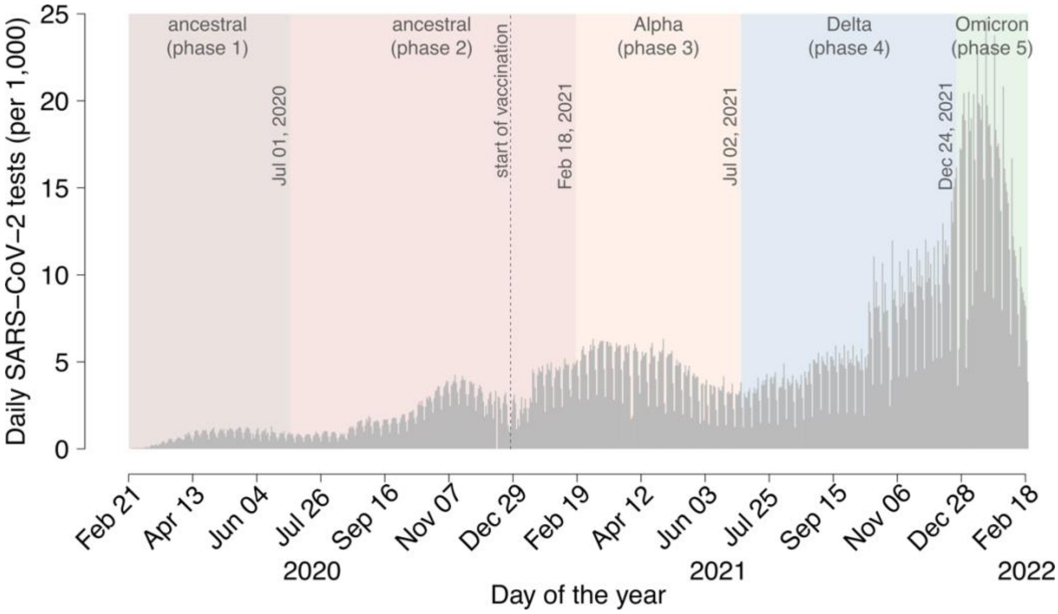

**Figure S5.** Grey bars represent the daily number of SARS-CoV-2 tests administered per 1,000 individuals (31). Background colors indicate the classification in different phases, and the dates indicated within the graph denote the day of transition between consecutive phases. The vertical dotted line denotes the start of the vaccination campaign on December 27, 2020.

Figure S6 shows the percentage of SARS-CoV-2 infections among vaccinated in Italy over time. This figure may assist the interpretation of the decrease in the ascertainment ratio found by our model since the Alpha phase. The shift of infection towards vaccinated population segments may have resulted in a higher amount of asymptomatic or pauci-symptomatic infections, which are more difficult to be detected by the surveillance system.

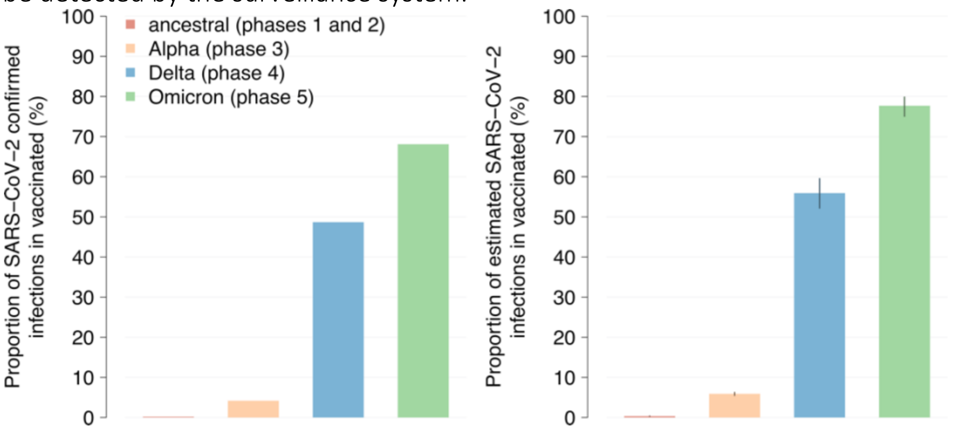

**Figure S6. a)** Percentage of SARS-CoV-2 confirmed infections reported to the national integrated surveillance system in vaccinated individuals (one or more doses, independently from the time at which vaccination was administered) in the ancestral phases (red), in the Alpha phase (orange), in the Delta phase (blue) and in the Omicron phase (green) (29). **b)** Proportion of SARS-CoV-2 infections in vaccinated individuals (one or more doses) as estimated by the model in the different phases. Bars: mean estimates; vertical lines: 95% CI; n=300 stochastic model realizations.

Figure S7 shows the estimated the distribution of the Italian population across groups characterized by different levels of susceptibility to SARS-CoV-2 infection at the end of simulations (February 20, 2020), considering the protection acquired from natural infection or vaccination and accounting for waning of both. Compared to Figure 3f in the main text, here we disaggregate the proportion of susceptible after waning of vaccination by the number of vaccine doses (two doses or including booster). Figure S7 suggests that waning of booster protection has begun to represent a significant proportion of the susceptible population, especially in older ages.

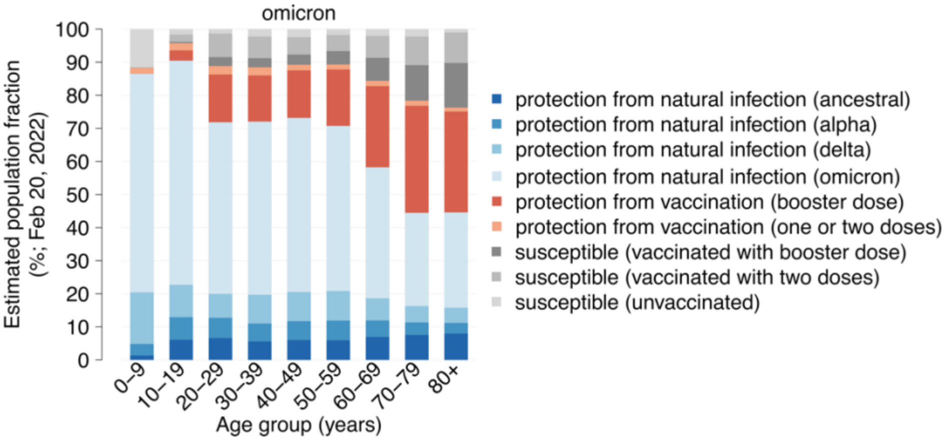

**Figure S7.** Mean estimates of the SARS-CoV-2 susceptibility profile by age groups at the end of simulations on February 20, 2022 (n=300 stochastic model realizations). In categories “protection from natural infection” the indicated variant corresponds to that of the most recent infection, independently of whether the individual has been vaccinated before or after natural infection. Uninfected and vaccinated Individuals whose immunity has not waned are included in the “protection from vaccination” categories, depending on whether they received or not a booster dose. Individuals who were never infected or whose immunity against infection (from natural infection or vaccine) has waned are considered in the “susceptible” categories depending on their vaccination status. Note that the latter may retain a residual protection against other clinical endpoints, such as symptoms, severe disease, or death.

### References

1. Marziano V, Guzzetta G, Mammone A, Riccardo F, Poletti P, Trentini F, et al. The effect of COVID-19 vaccination in Italy and perspectives for living with the virus. Nat Commun. 2021 Dec 14;12(1):7272.

2. Yang J, Marziano V, Deng X, Guzzetta G, Zhang J, Trentini F, et al. Despite vaccination, China needs non-pharmaceutical interventions to prevent widespread outbreaks of COVID-19 in 2021. Nat Hum Behav. 2021 Aug 1;5(8):1009–20.

3. Trentini F, Marziano V, Guzzetta G, Tirani M, Cereda D, Poletti P, et al. Pressure on the Health-Care System and Intensive Care Utilization During the COVID-19 Outbreak in the Lombardy Region of Italy: A Retrospective Observational Study in 43,538 Hospitalized Patients. Am J Epidemiol. 2022 Jan 1;191(1):137–46.

4. Guzzetta G, Riccardo F, Marziano V, Poletti P, Trentini F, Bella A, et al. Impact of a Nationwide Lockdown on SARS-CoV-2 Transmissibility, Italy. Emerg Infect Dis. 2020/10/20 ed. 2021 Jan;27(1):267–70.

5. Manica M, Guzzetta G, Riccardo F, Valenti A, Poletti P, Marziano V, et al. Impact of tiered restrictions on human activities and the epidemiology of the second wave of COVID-19 in Italy. Nat Commun. 2021 Jul 27;12(1):4570.

6. Istituto Superiore di Sanità. Monitoraggio delle varianti del virus SARS-CoV-2 di interesse in sanità pubblica in Italia [Internet]. [cited 2022 Apr 22]. Available from: https://www.epicentro.iss.it/coronavirus/sars-cov-2-monitoraggio-varianti-indagini-rapide

7. Mossong J, Hens N, Jit M, Beutels P, Auranen K, Mikolajczyk R, et al. Social Contacts and Mixing Patterns Relevant to the Spread of Infectious Diseases. PLOS Med. 2008 Mar 25;5(3):e74.

8. Hu S, Wang W, Wang Y, Litvinova M, Luo K, Ren L, et al. Infectivity, susceptibility, and risk factors associated with SARS-CoV-2 transmission under intensive contact tracing in Hunan, China. Nat Commun. 2021 Mar 9;12(1):1533.

9. Harris RJ, Hall JA, Zaidi A, Andrews NJ, Dunbar JK, Dabrera G. Effect of Vaccination on Household Transmission of SARS-CoV-2 in England. N Engl J Med. 2021 Aug 19;385(8):759–60.

10. Lipsitch M, Kahn R. Interpreting vaccine efficacy trial results for infection and transmission. Vaccine. 2021 Jul 5;39(30):4082–8.

11. Hall VJ, Foulkes S, Charlett A, Atti A, Monk EJM, Simmons R, et al. SARS-CoV-2 infection rates of antibody-positive compared with antibody-negative health-care workers in England: a large, multicentre, prospective cohort study (SIREN). The Lancet. 2021 Apr 17;397(10283):1459–69.

12. Altarawneh HN, Chemaitelly H, Hasan MR, Ayoub HH, Qassim S, AlMukdad S, et al. Protection against the Omicron Variant from Previous SARS-CoV-2 Infection. N Engl J Med. 2022 Mar 31;386(13):1288–90.

13. Fabiani M, Puopolo M, Morciano C, Spuri M, Alegiani SS, Filia A, et al. Effectiveness of mRNA vaccines and waning of protection against SARS-CoV-2 infection and severe covid-19 during predominant circulation of the delta variant in Italy: retrospective cohort study. BMJ. 2022 Feb 10;376:e069052.

14. Andrews N, Stowe J, Kirsebom F, Toffa S, Rickeard T, Gallagher E, et al. Covid-19 Vaccine Effectiveness against the Omicron (B.1.1.529) Variant. N Engl J Med. 2022 Apr 21;386(16):1532–46.

15. Fabiani M, Puopolo M, Filia A, Sacco C, Mateo-Urdiales A, Spila Alegiani S, et al. Effectiveness of an mRNA vaccine booster dose against SARS-CoV-2 infection and severe COVID-19 in persons aged ≥60 years and other high-risk groups during predominant circulation of the delta variant in Italy, 19 July to 12 December 2021. Expert Rev Vaccines. 2022 Apr 7;0(0):1–8.

16. Cereda D, Manica M, Tirani M, Rovida F, Demicheli V, Ajelli M, et al. The early phase of the COVID-19 epidemic in Lombardy, Italy. Epidemics. 2021 Dec 1;37:100528.

17. Manica M, Litvinova M, De Bellis A, Guzzetta G, Mancuso P, Vicentini M, et al. Estimation of the incubation period and generation time of SARS-CoV-2 Alpha and Delta variants from contact tracing data [Internet]. arXiv; 2022 Mar [cited 2022 May 23]. Report No.: arXiv:2203.07063. Available from: http://arxiv.org/abs/2203.07063

18. Manica M, De Bellis A, Guzzetta G, Mancuso P, Vicentini M, Venturelli F, et al. Intrinsic Generation Time of the SARS-CoV-2 Omicron Variant: An Observational Study of Household Transmission [Internet]. Rochester, NY: Social Science Research Network; 2022 Mar [cited 2022 Apr 26]. Report No.: 4068368. Available from: https://papers.ssrn.com/abstract=4068368

19. Covid-19 Opendata Vaccini [Internet]. Developers Italia; 2022 [cited 2022 Apr 12]. Available from: https://github.com/italia/covid19-opendata-vaccini

20. World Health Organization. WHO SAGE roadmap for prioritizing the use of COVID-19 vaccines in the context of limited supply: an approach to inform planning and subsequent recommendations based upon epidemiologic setting and vaccine supply scenarios, 13 November 2020 [Internet]. Version 1.1. Geneva: World Health Organization; 2020. Available from: https://apps.who.int/iris/handle/10665/341448

21. Dagan N, Barda N, Kepten E, Miron O, Perchik S, Katz MA, et al. BNT162b2 mRNA Covid-19 Vaccine in a Nationwide Mass Vaccination Setting. N Engl J Med. 2021 Apr 15;384(15):1412–23.

22. Polack FP, Thomas SJ, Kitchin N, Absalon J, Gurtman A, Lockhart S, et al. Safety and Efficacy of the BNT162b2 mRNA Covid-19 Vaccine. N Engl J Med. 2020 Dec 31;383(27):2603–15.

23. Diekmann O, Heesterbeek JAP, Metz JAJ. On the definition and the computation of the basic reproduction ratio R0 in models for infectious diseases in heterogeneous populations. J Math Biol. 1990 Jun 1;28(4):365–82.

24. Diekmann O, Heesterbeek J a. P, Roberts MG. The construction of next-generation matrices for compartmental epidemic models. J R Soc Interface. 2010 Jun 6;7(47):873–85.

25. Marziano V, Poletti P, Trentini F, Melegaro A, Ajelli M, Merler S. Parental vaccination to reduce measles immunity gaps in Italy. eLife. 2019 Sep 3;8:e44942.

26. Riccardo F, Ajelli M, Andrianou XD, Bella A, Manso MD, Fabiani M, et al. Epidemiological characteristics of COVID-19 cases and estimates of the reproductive numbers 1 month into the epidemic, Italy, 28 January to 31 March 2020. Eurosurveillance. 2020 Dec 10;25(49):2000790.

27. Task force COVID-19 del Dipartimento Malattie Infettive, Servizio di Informatica, Istituto Superiore di Sanità. Epidemia COVID-19. Aggiornamento nazionale: 2 marzo 2022 [Internet].

Available from: https://www.epicentro.iss.it/coronavirus/bollettino/Bollettino-sorveglianza-integrata-COVID-19_2-marzo-2022.pdf

28. Italian National Institute of Statistics (ISTAT). Popolazione residente al 1° Gennaio 2021 per età, sesso e stato civile [Internet]. [cited 2022 May 19]. Available from: https://demo.istat.it/popres/index.php?anno=2021&lingua=ita

29. Istituto Superiore di Sanità. COVID-19 ISS open data – EpiCentro. [Internet]. [cited 2022 Apr 19]. Available from: https://www.epicentro.iss.it/coronavirus/open-data/covid_19-iss.xlsx

30. Poletti P, Tirani M, Cereda D, Trentini F, Guzzetta G, Marziano V, et al. Age-specific SARS-CoV-2 infection fatality ratio and associated risk factors, Italy, February to April 2020. Eurosurveillance. 2020 Aug 6;25(31):2001383.

31. Dati COVID-19 Italia [Internet]. Presidenza del Consiglio dei Ministri - Dipartimento della Protezione Civile; 2022 [cited 2022 May 24]. Available from: https://github.com/pcm-dpc/COVID-19
